# Testing the association between tobacco and cannabis use and cognitive functioning: Findings from an observational and Mendelian randomization study

**DOI:** 10.1101/2020.02.26.20028274

**Authors:** Liam Mahedy, Robyn Wootton, Steph Suddell, Caroline Skirrow, Matt Field, Jon Heron, Matthew Hickman, Marcus R. Munafò

## Abstract

**Background:** Although studies have examined the association between tobacco and cannabis use in adolescence with subsequent cognitive functioning, study designs are usually not able to distinguish correlation from causation.

**Methods:** Separate patterns of tobacco and cannabis use were derived using longitudinal latent class analysis based on measures assessed on five occasions from age 13 to 18 in a large UK population cohort (ALSPAC). Cognitive functioning measures comprised of working memory, response inhibition, and emotion recognition assessed at 24 years of age. Mendelian randomization was used to examine the possible causal relationship.

**Results:** We found evidence of a relationship between tobacco and cannabis use and diminished cognitive functioning for each of the outcomes in the observational analyses. There was evidence to suggest that late-onset regular tobacco smokers (*b*=−0.29, 95%CI=−0.45 to −0.13), early-onset regular tobacco smokers (*b*=−0.45, 95%CI=−0.84 to −0.05), and early-onset regular cannabis users (*b*=−0.62, 95%CI=−0.93 to −0.31) showed poorer working memory. Early-onset regular tobacco smokers (*b*=0.18, 95%CI=0.07 to 0.28), and early-onset regular cannabis users (*b*=0.30, 95%CI=0.08 to 0.52) displayed poorer ability to inhibit responses. Late-onset regular (*b*=−0.02, 95%CI=−0.03 to −0.00), and early-onset regular tobacco smokers (*b*=−0.04, 95%CI=−0.08 to −0.01) showed poorer ability to recognise emotions. Mendelian randomization analyses were imprecise and did not provide additional support for the observational results.

**Conclusion:** There was some evidence to suggest that adolescent tobacco and cannabis use were associated with deficits in working memory, response inhibition and emotion recognition. Better powered genetic studies are required to determine whether these associations are causal.

## 1. INTRODUCTION

Tobacco and cannabis use during adolescence, when the brain is still developing and undergoing considerable structural and function changes (De Bellis et al., 2000), is a major public health concern. The association between adolescent tobacco and cannabis use and subsequent cognitive functioning has received particular attention because certain cognitive functions (e.g. working memory, response inhibition, and emotion recognition) do not peak until early adulthood (Davidson et al., 2006; Thomas et al., 2007) in parallel with maturation of the prefrontal cortex (Sowell et al., 2001). Due to the prolonged neurodevelopmental period and the potential for the endocannabinoid and nicotinic cholinergic signalling systems to be involved in altering development (Galve-Roperh et al., 2009; Newman and McGaughy, 2008), it is plausible that tobacco and cannabis use during this potentially critical period could play a role in disrupting normal brain development (Dwyer et al., 2008; Jacobus and Tapert, 2014). Nonetheless, there is still uncertainty regarding the nature of the association between tobacco and cannabis use and neurocognitive function.

A recent review of prospective studies of the association between cannabis use and cognition in young people (Gonzalez et al., 2017) highlighted an association between cannabis use and neuropsychological decline (Jackson et al., 2016). However, studies often fail to control for neurocognitive measures prior to cannabis use (Jacobus et al., 2015; Tait et al., 2011) and associations were largely found for the heaviest cannabis users and were often attenuated when potential confounders (e.g. other forms of substance use) were included (Jackson et al., 2016; Mokrysz et al., 2016b). A recent study (Meier et al., 2018), using a co-twin design (allowing for the disentanglement of shared genetic factors from non-shared environmental factors), assessed IQ prior to cannabis initiation and found insufficient evidence to suggest cannabis use was associated with decline in general IQ. Findings from two recent longitudinal studies of adolescents (Castellanos-Ryan et al., 2017; Morin et al., 2018) using a repeated measures design suggest that the association between cognitive functioning and cannabis use could be bidirectional.

The direction of association between tobacco and cognitive functioning is also unclear as there is a lack of epidemiological studies that have prospectively examined this relationship. Evidence from animal studies suggests that nicotine exposure may have more deleterious developmental effects during adolescence, when the brain is thought to be more vulnerable (Slotkin, 2002). Furthermore, human studies suggest that nicotine has a more potent effect when consumed in late adolescence compared to in adulthood (Azam et al., 2007). One small prospective study (*n*=112, aged between 17 to 21 years) found that current smokers performed worse than non-smokers on a variety of cognitive assessments including language related IQ and working memory while controlling for earlier cognitive measures and other substance use (Fried et al., 2006). Finally, one large study (*n*∼20,000) on Israeli male soldiers (Weiser et al., 2010) found a dose-response relationship between number of cigarettes smoked and lower general cognitive ability compared to non-smokers. They also found diminished cognitive functioning in individuals who started starting smoking after 18 years of age. The literature is further complicated by the differential effects of acute, chronic, and withdrawal from chronic nicotine on cognitive functioning. Studies have reported beneficial effects of acute nicotine (Heishman et al., 2010), negative effects of nicotine withdrawal on cognitive functioning (Mendrek et al., 2006), and the reduction of beneficial effects with nonacute nicotine consumption as tolerance develops (Jacobsen et al., 2005).

In an effort to strengthen the evidence, we used data from the Avon Longitudinal Study of Parents and Children (ALSPAC), a large UK prospective birth cohort, to investigate whether patterns of adolescent tobacco and cannabis use were prospectively associated with cognitive functioning at 24 years of age. Separate measures of tobacco and cannabis use were assessed on six occasions across adolescence allowing distinct classes of tobacco and cannabis use to be established. As young people do not initiate tobacco or cannabis at the same time (Degenhardt et al., 2008), we used longitudinal latent class analysis to identify heterogenous classes of individuals with different tobacco and cannabis use profiles across adolescence (Lubke and Muthén, 2005). As a next step we used genetic variants that are separately associated with smoking initiation and lifetime cannabis use to perform Mendelian randomization (MR) to improve causal inference (Lawlor et al., 2017). The aims were to investigate (1) whether separate patterns of tobacco smoking and cannabis use (assessed between 13 to 18 years) were associated with working memory, response inhibition, and emotion recognition assessed at age 24, and (2) whether tobacco use and cannabis use were associated with these cognitive outcomes using MR.

## 2. METHODS

### 2.1 Observational analyses

#### 2.1.1 Participants and Procedure

The Avon Longitudinal Study of Parents and Children (ALSPAC) is a cohort born in 1991– 92. ALSPAC recruited 14,541 pregnant women resident in Avon, UK, with expected dates of delivery between 1 April 1991 and 31 December 1992. The initial number of pregnancies enrolled is 14,541 (for these at least one questionnaire has been returned or a “Children in Focus” clinic had been attended by 19/07/99). Of these initial pregnancies, there was a total of 14,676 foetuses, resulting in 14,062 live births and 13,988 children who were alive at 1 year of age. When the oldest children were approximately 7 years of age, an attempt was made to bolster the initial sample with eligible cases who had failed to join the study originally. The total sample size for analyses using any data collected after the age of 7 years is therefore 15,454 pregnancies, resulting in 15,589 foetuses. Of this total sample 14,901 were alive at 1 year of age (Boyd et al., 2013; Fraser et al., 2013; Northstone et al., 2019). Of these, 9,997 offspring were invited to attend the 24-year clinic assessment. A detailed overview of our study population, including attrition at the different measurement occasions is presented in Supplementary Material Figure S1. Detailed information about ALSPAC is available online www.bris.ac.uk/alspac. A fully searchable data dictionary is available on the study’s website http://www.bristol.ac.uk/alspac/researchers/our-data/. Approval for the study was obtained from the ALSPAC Ethics and Law Committee and the Local Research Ethics Committees (http://www.bristol.ac.uk/alspac/researchers/research-ethics/). Informed consent for the use of data collected via questionnaires and clinics was obtained from participants following recommendations of the ALSPAC Ethics and Law Committee at the time. Consent for biological samples was collected in accordance with the Human Tissue Act (2004).

### 2.2 Measures

A timeline of data collection is presented in Supplementary Material Figure S2.

#### 2.2.1 Exposure variables

Information on tobacco and cannabis use were collected on six occasions via questionnaire (Q) or during attendance at a study clinic (C). Median ages at response were: 13y(C), 14y(Q), 15y(C) 16y(Q), 17y(C), and 18y(Q).

#### 2.2.2 Tobacco use

Patterns of tobacco use have been described in detail elsewhere (Howe et al., 2017). Responses to one or more questions at each time point were used to derive a repeated four-level ordinal variable with categories ‘Non-smoker’, ‘Occasional smoker’ (typically less than once per week), ‘Weekly smoker’ and ‘Daily smoker’. There was good agreement that a four-class solution was adequate in explaining the heterogeneity in tobacco based on model fit criteria (see Table S1a). The four-class model (*n*=8,525) comprised individuals with a higher probability of ‘early-onset regular tobacco smokers’ (3.4%), ‘late-onset regular tobacco smokers’ (11.6%), ‘experimenters’ (17.4%), and ‘non-tobacco smokers’ (67.5%) (Figure S3a).

#### 2.2.3 Cannabis use

Patterns of cannabis use have been described in detail elsewhere (Taylor et al., 2017). Responses to one or more questions at each time point were used to derive a repeated three-level ordinal variable with categories ‘Do not use’, ‘Occasional use’ (typically less than once per week) and ‘Frequent use’ (typically once per week or more). There was good agreement that a four-class solution was adequate in explaining the heterogeneity in cannabis use based on model fit criteria (see Table S1b). The four-class model (*n*=8,093) comprised individuals with a higher probability of ‘early-onset regular users’ (3.6%), ‘early-onset occasional users’ (2.9%), ‘late-onset occasional users’ (13.8%), and ‘non-users’ (79.8%) (Figure S3b).

#### 2.2.4 Outcome variables

At 24 years of age (*M*=24.0 years; *SD*=9.8 months) participants attended a clinic-based assessment which included computerised cognitive assessments as part of a broader assessment battery of mental and physical health and behaviour. Data collection for the online questionnaires was collected and managed by REDcap electronic data capture tools (Harris et al., 2019, 2009). Further information on all three cognitive tasks is presented in Supplementary material.

#### 2.2.5 Working memory

The *N*-back task (2-back condition) was used to assess working memory. The *N*-back task (Kirchner, 1958) is widely used to measure working memory. A measure of discriminability (*d*′) was chosen as the primary outcome measure given it is an overall performance estimate. High scores on *number of hits* indicating more accurate identification, while high scores on *false alarms* indicating less accurate identification were examined as secondary outcomes. High scores on *d*′, therefore, indicated a greater ability to distinguish signal from noise. *d*′ data were available for *n*=3,242 participants.

#### 2.2.6 Response inhibition

The Stop Signal Task (Logan et al., 1984) was used to assess response inhibition – the ability to prevent an ongoing motor response. The task consisted of 256 trials, which included a 4:1 ratio of trials without stop signals to trials with stop signals. Mean response times were calculated. An estimate of *stop signal reaction time* (SSRT) was calculated using the median of the inhibition function approach (Band et al., 2003). SSRT used as the primary outcome as it is a reliable measure of inhibitory control, with shorter reaction times indicating faster inhibition. SSRT data were available for 3,201 participants. Individual Stop Signal indices (i.e., Go reaction time, Stop accuracy, and Go accuracy) were examined as secondary outcomes.

#### 2.2.7 Emotion recognition

Emotion recognition was assessed using a six alternative forced choice (6AFC) emotion recognition task (Penton-Voak et al., 2012) comprising of 96 trials (16 for each emotion) which measures the ability to identify emotions in facial expressions that vary in intensity. In each trial, participants were presented with a face displaying one of six emotions: anger, disgust, fear, happiness, sadness, or surprise. Participants were required to select the descriptor that best described the emotion that was present in the face, using the computer mouse. Emotion intensity varied across 8 levels within each emotion from the prototypical emotion to an almost neutral face. Each individual stimulus was presented twice, giving a total of 96 trials. An overall measure of emotion recognition (the number of facial emotions accurately identified) was used as the primary outcome. Emotion recognition data were available for *n*=3,368 participants. Each of the individual emotions were examined as secondary outcomes.

#### 2.2.8 Potential confounders

Confounders comprised of established risk factors for cognitive functioning that could plausibly have a relationship with earlier substance use. Potential confounders included: income, maternal education, socioeconomic position, housing tenure, sex, and maternal smoking during first trimester in pregnancy. Working memory at approximately 11 years and experience of a head injury/unconsciousness up to 11 years were included to control for cognitive functioning prior to baseline measures of substance use. Finally, a measure of alcohol use asking whether they had ever had a whole drink of alcohol was collected at age 13 years (up to the first assessment of smoking and cannabis use). Further information is presented in Supplementary Material.

### 2.3 Statistical methods

Different tobacco and cannabis phenotypes were used across different analytic methods.

#### 2.3.1 Observational analyses

Tobacco and cannabis class membership was related to covariates using the Bolck-Croon-Hagenaars (Bolck et al., 2004) method. This approach uses the weights derived from the latent classes to reflect measurement error in the latent class variable. Linear regression was used to examine the association between the cognitive outcomes and latent class membership controlling for the confounding variables. Results are reported as unstandardized beta coefficients with 95% confidence intervals. Analyses were carried out using Mplus 8.4 (Muthén and Muthén, 2016).

#### 2.3.2 Missing data

Missing data was dealt with in three steps. First, full information maximum likelihood (FIML) was used to derive trajectories tobacco (*n*=8,525) and cannabis (*n*=8,093) based on individuals who had information on at least one timepoint between 13 and 18 years. For a detailed description of missingness at each timepoint see Tables S2a and S2b. Next, multiple imputation was based on 3,232 participants (for both tobacco and cannabis models) who had information on at least one of the cognitive outcomes. The imputation model (based on 100 datasets) contained performance on all of the cognitive tasks, all measures of tobacco and cannabis use, and potential confounding variables, as well as a number of auxiliary variables known to be related to missingness (e.g., substance use in early adolescence, parental financial difficulties, and other SES variables). Finally, inverse probability weighting was used where estimates of prevalence and associations were weighted to account for probabilities of non-response to attending the clinic. See Table S3 for a detailed description of attrition for completing the cognitive assessments at age 24 years. See Tables S4a and S4b for a detailed description of confounding factors associated with tobacco and cannabis use class membership. See Table S5 for a detailed description of sample characteristics.

#### 2.3.3 Genetic analyses

Our aim was to triangulate the findings from the observational analyses with one- and two-sample MR analyses. However, due to insufficient power in the two-sample MR analyses, we will primarily focus on the one-sample MR results. Two-sample MR are still included as a set of sensitivity analyses as they allow us to conduct some of the pleiotropy robust methods (e.g., MR-Egger, weighted median, and weighted mode), but must be interpreted with caution. Information on genotyping and quality control are presented in the Supplementary Material.

#### 2.3.4 Mendelian randomization (MR)

One-sample MR analyses using two-stage least squares regression models with robust standard errors was used to examine the to examine the polygenic risk score constructed using genome-wide significant SNPs for smoking initiation (378 SNPs (Liu et al., 2019) as an instrument for smoking initiation and cannabis use (8 SNPs (Pasman et al., 2018) as an instrument for lifetime cannabis use in relation to the three cognitive assessments at 24 years of age. Using individual-level data, the first stage involves regressing tobacco/cannabis use upon SNPs for individual smoking initiation/lifetime cannabis use. Lifetime tobacco use (*n*=1,638/5,107) (32%) up to age 15 years and lifetime cannabis use (*n*=1,348/5,319) (25%) up to age 24 years were chosen as exposures. Each of the cognitive outcomes were then regressed on the fitted values from the stage 1 for tobacco and cannabis use in the second stage. The three key assumptions in MR are 1) the genetic instrument is robustly associated with the exposure of interest; 2) confounders of the exposure-outcome association are not associated with the genetic instrument; and 3) the genetic instrument is not associated with the outcome other than through its association with the exposure; see (Lawlor et al., 2017) for a full description. Power calculations conducted for one-sample MR analyses using mRnd (Brion et al., 2012) indicated that we had 80% power to detect an effect size of 0.24 for smoking initiation and 0.15 for lifetime cannabis use using a sample size of *n*∼3,300 (individuals who had available cognitive data in ALSPAC).

#### 2.3.5 Sensitivity analyses

Two-sample MR analysis was used to test the hypothesised causal effect of smoking initiation and lifetime cannabis use on cognitive functioning. See Supplementary material for further details.

## 3. RESULTS

### 3.1 Observational analyses

#### 3.1.1 Patterns of tobacco use

Fully adjusted associations between patterns of tobacco use from 13 to 18 years and cognitive functioning outcomes at age 24 are presented in Table 1. There was evidence to suggest that late-onset regular tobacco smokers demonstrated poorer working memory performance (*b*=−0.29, 95%CI=−0.45 to −0.13) and emotion recognition (*b*=−0.02, 95%CI=−0.03 to −0.00) compared to non-tobacco users. While early-onset regular tobacco smokers showed poorer performance across all three cognitive outcomes compared to the non-tobacco users: working memory (*b*=−0.45, 95%CI=−0.84 to −0.05), response inhibition (*b*=0.18, 95%CI=0.07 to 0.28), and emotion recognition (*b*=−0.04, 95%CI=−0.08 to −0.01). All associations were supported by significant Wald test values indicating a significant difference between the groups. Results demonstrating various levels of adjustment are presented in the Supplementary Material (Tables S6a-S6c).

**Table 1.** Smoking patterns from 13 to 18 years and cognitive functioning at age 24 (fully adjusted models)

|  | No smoking | Experimenter | Late-onset regular | Early-onset regular |  |
| --- | --- | --- | --- | --- | --- |
| *n*=3,232 for all models | Reference | *b* (95% CI) | *b* (95% CI) | *b* (95% CI) | Wald (df) *p* value |
| **Working memory** | - | 0.01 (-0.12, 0.10) | -0.29 (-0.45, -0.13) | -0.45 (-0.84, -0.05) | 22.12 (3) *p*<0.001 |
| **Response inhibition** | - | 0.01 (-0.06, 0.09) | 0.10 (-0.12, 0.32) | 0.18 (0.07, 0.28) | 12.78 (3) *p*=0.005 |
| **Emotion recognition** | - | -0.00 (-0.01, 0.01) | -0.02 (-0.03, -0.00) | -0.04 (-0.08, -0.01) | 16.43 (3) *p*=0.001 |

Note. Models adjusted for socioeconomic status, working memory at age ~11 years; head injury/ unconsciousness up to age 11 years, and alcohol use before 13 years of age; Wald tests determine whether there were differences between patterns of tobacco use and subsequent cognitive functioning; Working memory: negative *d'* scores reflect poorer performance; Response inhibition: longer reaction times reflect poorer performance; Emotion recognition: negative scores reflect poorer performance

In the secondary analyses, there was some evidence to suggest that early-onset regular tobacco smoking was associated with fewer correct hits on the *N*-back task in the fully adjusted models (Table S7). There was evidence to suggest that late-onset regular tobacco users were associated with poorer Go and Stop accuracy in the fully adjusted models (Table S8). There was some evidence to suggest late-onset regular tobacco users had poorer ability to identify ‘fear’ and ‘surprise’, while early-onset regular tobacco users had poorer ability to identify ‘sad’ in the fully adjusted models (Table S9d).

#### 3.1.2 Patterns of cannabis use

Fully adjusted associations between patterns of cannabis use from 13 to 18 years and cognitive functioning outcomes at age 24 are presented in Table 2. There was evidence to suggest that early-onset regular cannabis users showed poorer working memory performance (*b*=−0.62, 95%CI=−0.93 to −0.31) and response inhibition (*b*=0.30, 95%CI=0.08 to 0.52) compared to non-cannabis users. All associations were supported by significant Wald test values indicating a significant difference between the groups. Results demonstrating various levels of adjustment are presented in the Supplementary Material (Tables S10a-10c).

**Table 2.** Patterns of cannabis use from 13 to 18 years and cognitive functioning at age 24 (fully adjusted models)

|  | Non-user | Late-onset occasional | Early-onset occasional | Early-onset regular |  |
| --- | --- | --- | --- | --- | --- |
| *n*=3,232 for all models | Reference | *b* (95% CI) | *b* (95% CI) | *b* (95% CI) | Wald (df) *p* value |
| **Working memory** | - | -0.10 (-0.22, 0.03) | 0.12 (-0.17, 0.41) | -0.62 (-0.93, -0.31) | 18.56 (3) *p*<0.001 |
| **Response inhibition** | - | 0.04 (-0.04, 0.11) | 0.05 (-0.11, 0.22) | 0.30 (0.08, 0.52) | 9.24 (3) *p*=0.02 |
| **Emotion recognition** | - | -0.00 (-0.01, 0.01) | 0.02 (-0.00, 0.05) | -0.02 (-0.05, 0.01) | 4.23 (3) *p*=0.24 |

Note. Models adjusted for socioeconomic status, working memory at age ~11 years; head injury/ unconsciousness up to age 11 years, and alcohol use before 13 years of age; Wald tests determine whether there were differences between patterns of tobacco use and subsequent cognitive functioning; Working memory: negative *d'* scores reflect poorer performance; Response inhibition: longer reaction times reflect poorer performance; Emotion recognition: negative scores reflect poorer performance

In the secondary analyses, there was evidence to suggest that early-onset cannabis users were associated with worse Go and Stop accuracy compared to non-cannabis users, in the fully adjusted models (Table S11). There was no evidence of an association between specific response inhibition measures in the sensitivity analyses (Table S12). Early- and late-onset occasional cannabis users had better ability to identify ‘anger’ and ‘disgust’, while late-onset occasional cannabis users had poorer ability to identify ‘happy’ in the fully adjusted models (Table 13d).

### 3.2 Genetic analyses

Information testing whether the genetic instruments are associated with the confounders are presented in the Supplementary Material (Tables S14a and S14b).

#### 3.2.1 One-sample Mendelian randomization

Results from the one-sample MR provided little evidence to suggest that smoking initiation or lifetime cannabis use were causal risk factors for deficits in cognitive functioning (Table 3). SNPs associated with smoking initiation were not associated with working memory (*b*=−0.38 95%CI=−1.51 to 0.75; *p*=0.51); response inhibition (*b*=−0.27 95%CI=−1.31 to 0.77; *p*=0.61); or emotion recognition (*b*=−0.53 95%CI=−1.59 to 0.53; *p*=0.33). Similarly, SNPs associated with lifetime cannabis use were not associated with working memory (*b*=−1.41 95%CI=−3.31 to 0.49; *p*=0.14); response inhibition (*b*=0.02 95%CI=−1.31 to 1.35; *p*=0.98); or emotion recognition (*b*=−0.04 95%CI=−1.35 to 1.27; *p*=0.95).

**Table 3.** One-sample MR analyses of the effects of smoking initiation on cognitive functioning (standardised coefficients)

| N=1,638 | $\beta$ | se | 95% CI | *p* | F statistic |
| --- | --- | --- | --- | --- | --- |
| **Smoking initiation** |  |  |  |  |  |
| Working memory | -0.38 | 0.58 | -1.51, 0.75 | 0.51 | 14.63 |
| Response inhibition | -0.27 | 0.53 | -1.31, 0.77 | 0.61 | 18.12 |
| Emotion recognition | -0.53 | 0.54 | -1.59, 0.53 | 0.33 | 17.57 |
| **Lifetime cannabis use** |  |  |  |  |  |
| N=1,348 |  |  |  |  |  |
| Working memory | -1.41 | 0.97 | -3.31, 0.49 | 0.14 | 6.70 |
| Response inhibition | 0.02 | 0.68 | -1.31, 1.35 | 0.98 | 8.87 |
| Emotion recognition | -0.04 | 0.69 | -1.35, 1.27 | 0.95 | 8.79 |

#### 3.2.2 Sensitivity analyses

Overall, the two-sample MR methods provided some evidence to suggest that SNPs associated with smoking initiation and SNPs associated with lifetime cannabis use were a causal risk factor for deficits in cognitive functioning. See Supplementary Material (Tables S15a and S15b).

## 4. DISCUSSION

This observational study provided evidence to suggest an association between tobacco and cannabis use across adolescence and subsequent cognitive functioning. Early- and late-onset regular tobacco smokers demonstrated poorer working memory and poorer ability to recognise emotions; while, early-onset regular tobacco smokers had slower ability to inhibit responses compared to non-tobacco smokers. Early-onset regular cannabis users had poorer working memory performance and slower ability to inhibit responses compared to non-cannabis users. Our results remained largely consistent when controlling for prior measures of substance use and cognition allowing for clear temporality between exposure and outcomes. Genetic analyses were imprecise and did not provide sufficient evidence for a possible causal association between smoking initiation and lifetime cannabis use and cognitive functioning in the ALSPAC sample. It is likely that these analyses were underpowered.

### 4.1 Comparison with previous studies

To our knowledge, this is the first study to assess the relationship between separate tobacco and cannabis use in adolescents, and subsequent cognitive functioning using a combination of observational and genetic epidemiological approaches. Overall, we found an adverse association between tobacco/cannabis use and working memory, response inhibition, and emotion recognition in ALSPAC. Those who initiated regular use at earlier and later ages demonstrated poorer performance on the cognitive tasks. There was some evidence to suggest cannabis use with associated with emotion-specific impairments in emotion recognition. This is in line with previous research suggesting cannabis users may have poorer recognition of negative emotions (Bossong et al., 2013). Our results also tentatively suggest that recognition deficits may be related to specific patterns of cannabis use, with different patterns in early- and late-onset use. The observational findings contribute to a literature of mixed findings regarding the direction of association between tobacco and cannabis exposure and subsequent cognition by suggesting that adolescent tobacco and cannabis use precede observed reductions in cognitive function. These findings support studies that have demonstrated effects may depend on the frequency, duration, and age at onset of use (Boccio and Beaver, 2017; Castellanos-Ryan et al., 2017; Fontes et al., 2011; Mashhoon et al., 2018; Meier et al., 2018; Mokrysz et al., 2016a).

Our study extends previous findings in a number of ways. First, the observational study was better powered than most of the previous studies as it used data from over 3,200 participants providing information spanning birth to 24 years of age. Second, identifying heterogeneous patterns of tobacco and cannabis use across this crucial period allows individuals who follow markedly different developmental trajectories to be captured (Chen and Kandel, 1995; Degenhardt et al., 2008). Third, the cognitive measures were assessed at a time when they are expected to have reached maturity in some individuals (Davidson et al., 2006; Fry and Hale, 2000; Kramer et al., 2005; Thomas et al., 2007), in comparison to previous studies which have examined cognitive functioning at earlier ages while they are still maturing. Examining mature levels of cognitive functioning reduces the possibility that cognitive functioning is influencing earlier tobacco and cannabis use, effects that cannot be disentangled in purely cross-sectional studies. Further, our ability to control for earlier measures of cognitive functioning and substance use, prior to the baseline measures of tobacco and cannabis use helps to rule out the possibility of reverse causation. Fourth, our study sought to examine specificity in cognitive functioning, by using well-validated tests to probe different domains of cognitive functioning instead of focusing on general intelligence. Finally, we sought to triangulate our results by using one- and two-sample MR approaches to assess tobacco and cannabis use as causal risk factors for cognitive functioning. This approach can help to overcome the main sources of bias from classical observational approaches, by providing a more reliable estimate of the likely underlying causal relationship.

### 4.2 Limitations

There are limitations to this study that should be considered. First, the ALSPAC cohort suffers from attrition, which is higher among the socially disadvantaged (Wolke et al., 2009). Furthermore, polygenic scores for tobacco smoking initiation were associated with drop out in the ALSPAC (Taylor et al., 2018). We attempted to minimize the effect of drop-out by using multiple imputation, FIML, and inverse probability weighting which assume MAR missing patterns. Although it is not possible to test the MAR assumption, it was made more plausible as a number of SES variables were found to predict whether participants attended the clinic or not (Table S1). Second, tobacco and cannabis use were self-reported. However, there is evidence to suggest that self-reported assessments are reliable and valid methods (Boykan et al., 2019), and the assessment of tobacco and cannabis use yearly over 6 years in a latent variable framework helps to account for measurement error (Bray et al., 2015). Third, while the longitudinal approach for each substance used in this study has a number of advantages over using measures at a single timepoint, it was not possible to examine cannabis use without tobacco use as most cannabis users use cannabis in combination with tobacco (Amos et al., 2004). We therefore cannot rule out the possibility that observed associations between cannabis use and cognitive functioning are exacerbated by the combined use of cannabis and tobacco. Fourth, different measures of tobacco and cannabis use for the observational and MR analyses were used. Along with deriving latent classes of tobacco and cannabis use, we used the largest GWAS consortia (GSCAN) which has identified 341 genetic instruments for ‘smoking initiation’, and the GWAS conducted by Pasman and colleagues which identified 8 genetic instruments for lifetime cannabis use which are continuous measures. To our knowledge it is not currently possible to use a nominal exposure (as was used in the observational analyses) and consequently the effect sizes are not directly comparable. Fifth, it is likely that both the one- and two-sample MR analyses are underpowered. However, findings using weak instruments tend to bias findings towards the null in the two-sample setting and toward the outcome-risk association in the one-sample setting (Davies et al., 2018). Sixth, the main limitation of one- and two-sample MR is that the quality of the pooled results in the GWAS consortia is dependent on the individual studies. Another limitation is that the same sample may contribute to both GWAS (i.e., GWAS for exposure and outcome) which was the case in the current study as ALSPAC was in both the exposure and outcome. This will bias the MR estimate towards the observed estimate. However, as the MR found no clear evidence for an effect, this suggests it was not biased by overlapping samples. See Lawlor and colleagues (Lawlor et al., 2017) for a more comprehensive description of limitations associated with MR studies. Finally, it is possible that the direction of the association could work in both ways, that is, impairments in cognitive functioning may precede (and increase the risk of developing) tobacco and cannabis use (Anokhin and Golosheykin, 2016; Castellanos-Ryan et al., 2017; Cousijn et al., 2014). We were able to include a number of measures to maximize the robustness of our findings: (i) ascertaining the temporal order of exposures and outcomes; (ii) controlling for premorbid working memory and brain insults prior to measures of tobacco/cannabis use helped to reduce the possibility of cognitive impairments, or lower cognitive abilities in childhood, influencing tobacco/cannabis use; and (iii) it is possible that a common risk factor is influencing both tobacco/cannabis use and lower cognitive function, however MR methods helps to protect against this possibility by minimizing bias from reverse causation and residual confounding.

### 4.3 Implications and conclusions

Overall, there was observational evidence that adolescent tobacco and cannabis use were associated with subsequent cognitive functioning, highlighting impairments in a range of cognitive domains, including working memory, response inhibition and emotion recognition. Our findings lend support to the developmental vulnerability hypothesis, in that, tobacco and cannabis use in adolescence, when the brain is undergoing critical development, may have neurotoxic effects. Better powered genetically informed studies are required to determine whether these associations are causal. In order to rule out the possibility of deficient cognitive functioning preceding substance in adolescence, future research should use an equally robust approach to examine the alternate hypothesis. This study lends support to public health strategies and interventions aimed at reducing tobacco and cannabis exposure in young people.

## Data Availability

We used data from the Avon Longitudinal Study of Parents and Children (ALSPAC), an ongoing population-based study that contains a wide range of phenotypic and environmental measures, genetic information and linkage to health and administrative records.

http://www.bristol.ac.uk/alspac/researchers/our-data/

http://www.bris.ac.uk/alspac

## Contributors

This publication is the work of the authors and LM, MH, JH, and MRM will serve as guarantors for the contents of this paper. All authors have participated in the preparation of the manuscript and approve of its submission. **Conceptualization:** LM, MH, JH, and MRM; **Formal analysis:** LM, REW, SS, and CS; **Funding acquisition:** MH, JH, MF, and MRM; **Methodology:** LM, JH, and MRM; **Writing – original draft:** LM; **Writing – review & editing:** LM, REW, SS, CS, MF, MH, JH, and MRM. The views expressed in this publication are those of the authors and not necessarily those of the NHS, the National Institute for Health Research or the Department of Health and Social Care.

## Acknowledgements

We are extremely grateful to all the families who took part in this study, the midwives for their help in recruiting them, and the whole ALSPAC team, which includes interviewers, computer and laboratory technicians, clerical workers, research scientists, volunteers, managers, receptionists and nurses. GWAS data were generated by Sample Logistics and Genotyping Facilities at Wellcome Sanger Institute and LabCorp (Laboratory Corporation of America) using support from 23andMe. A comprehensive list of grants funding is available on the ALSPAC website (http://www.bristol.ac.uk/alspac/external/documents/grant-acknowledgements.pdf).

