## Supplementary material for "Testing the association between tobacco and cannabis use and cognitive functioning: Findings from an observational and Mendelian randomization study": tob and can_SM_MedRxiv.pdf

*Cognitive functioning measures*

*Working memory.* Participants continuously monitored a series of numbers presented on a computer screen and pressed '1' if the number was the same as the number presented *N* numbers ago, or '2' if it was not. Stimuli were numbers 0–9, presented in black on white background with a random spatial jitter of 180 pixels in y-axis and 200 pixels in x-axis. Each target was presented for 500 ms, followed by a 3,000 ms response window. The practice block consisted of 12 trials containing two targets. The experimental block consisted of 48 trials, containing 8 targets, where the target was the number that was identical to the one presented 2 trials back. Three outcomes were examined for the *N*-back task (i) number of hits, or the percentage of matching numbers correctly identified as matches, (ii) false alarms, or the percentage of non-matching numbers incorrectly identified as matches, and (iii) discriminability index, *d′*, which is a signal-detection metric that takes into account both hits and false alarms to derive an overall estimate of signal-detection ability (McNicol, 1972). *d′* was calculated using the Stata syntax adapted from (Stanislaw & Todorov, 1999). Of the participants assessed with cognitive tasks at age 24 (*n*=3,312), *n*=182 did not provide any data on the task; *n*=70 were omitted due to negative *d'* scores and/or not responding to over 50% of the trials.

$$d' = invnorm(\text{hits}) - invnorm(\text{false alarms})$$

*Response inhibition.* Participants were asked to sit in front of a computer monitor and their two index fingers were placed in two stimulus boxes, one labelled X and one labelled O. Two types of trials were performed, 'go' trials and stop signal trials. In the 'go' trials, participants were asked to fixate on a plus sign (+) in the centre of the computer screen. An X or O was presented on the screen and the participant had to press the corresponding button as quickly as possible. On 25% of the trials, a beep is heard (stop signal), randomly after the X and O appears. Participants were told to not press a response button when the beep was sounded, and to wait for the next trial to begin. If the beep was not heard the participant was asked to press the corresponding key according to what was presented on screen. When the beep was sounded, the participant was to refrain from pressing the response button.

32 practice trials were presented. The task consisted of 256 trials, comprised of 4 blocks of 64 trials. Each block of 64 trials consists of 4 sub-blocks of 16 trials. Each sub-block consists of 12 trials without a stop-signal and 4 trials with a stop-signal. Mean response times were calculated. Four metrics were examined for the stop signal task: (i) an estimate of stop signal reaction (SSRT) was calculated and used as the primary outcome as it is a reliable measure of inhibitory control, with shorter SSRT's indicating slower inhibition; secondary outcomes include: (ii) 'go' reaction time; (iii) 'go' accuracy; and (iv) 'stop' accuracy.

$$\text{SSRT}_{\text{med}} = \text{Go Reaction Time}_{\text{med}} - \text{Stop Signal Delay}_{\text{med}}$$

Stop Signal $\text{Delay}_{\text{med}}$ (SSD) was calculated for each session using a weighted least squares linear regression to predict SSD based on the probability of responding given a stop-signal. This was then used to estimate the SSD where the probability of the participant failing to inhibit was 50%.

*Emotion recognition.* Prototypical composite images of the six basic facial expressions of emotion were generated from 12 individual male faces showing each of the six expressions. The 12 original images were each delineated with 172 feature points, which allowed both shape and colour information to be averaged across the faces to generate ‘average’ anger, sadness, surprise, disgust, fear, happiness, using established techniques. An overall emotional prototype face was then generated by averaging the exemplars for each emotional expression. Facial images showing a specific emotion were displayed on the screen one at a time. Images were presented for 200 ms, followed by a backwards mask (white noise) of 250 ms. Participants were required to select the descriptor that best described the emotion that was present in the face, using the computer mouse. Emotion intensity is varied across 8 stimuli within each emotion on a scale from the most prototypical emotion to an almost neutral emotion. Each individual stimulus is presented twice, giving a total of 96 trials. The task was delivered using E-Prime Professional v. 2.0 software (Schneider, Eschman, & Zuccolotto, 2002).

For each of the specific emotions an unbiased hit rate was derived and used as the secondary outcome. This is based on work by Wagner (Wagner, 1993) who proposed an alternative score, the “unbiased hit rate” ($H_u$), designed to account for response biases. $H_u$ for each participant is calculated as the squared frequency of correct responses for a target emotion divided by the product of the number of stimuli representing this emotion and the overall frequency of this emotion category being chosen. $H_u$ has a range of zero to one, one indicating that all stimuli of an emotion have been correctly identified and the respective emotion has never been falsely chosen for a different emotion. Results from the secondary analyses are presented in Tables S8a-d.

*Potential confounders*

Confounders included: income (quintiles), maternal education (<O level: indicating no qualification; O level: indicating completion of school examinations at age 16; and >O level: indicating completion of college or university education at or after age 18), socioeconomic position (SEP, grouped into four categories: (a) unskilled or semiskilled manual; (b) skilled manual or non-manual; (c) managerial and technical and (d) professional), housing tenure (mortgaged, subsidised renting and private renting), sex, and maternal smoking during first trimester in pregnancy (yes/no).

A computerized version of the Counting Span task (Case, Kurland, & Goldberg, 1982) was included at approximately 11 years (*M*=10 years 8 months, SD=3 months) to assess working memory performance during a clinic visit. A span score was based on the number of correctly recalled sets (maximum score of 5 in increments of 0.5). Since adolescents who have experienced head injury perform poorly in working memory tasks compared with age-matched peers (Newsome et al., 2007), we covaried for head injury/unconsciousness before the age of 11, *n*=113 (3.4%). Finally, prior substance use was assessed using ever having consumed alcohol or used tobacco before 13 years of age (yes/no).

*Missing data*

Missing data on cannabis and tobacco use were dealt with using full information maximum likelihood. SES confounders assessed largely in pregnancy had minimal missing data (e.g., parental social class had the most missing data: 1,069/8,093 (13%), while the cognitive measures assessed up to age 11 years and substance use assessed at age 16.5 years had moderate missing data 2,034/8,093 (25%). Given that the BCH method uses listwise deletion for the outcome measures, *n*=2,073 participants had complete information on cannabis use, 2,059 had complete information on tobacco use and outcome and confounder data and at least one measure of tobacco use.

*Inverse probability weighting*

Weights were derived from logistic regression models using variables associated with nonresponse, including maternal age, grandmother having a history of severe depression, maternal alcohol use in pregnancy, financial problems, maternal cannabis use and financial problems. We weighted the included respondents by the inverse of the probability of attending and used the Hosmer-Lemeshow test to assess model fit.

*Model fit LLCA of tobacco and cannabis use*

Criteria for best fit included i) information-theoretic methods with lower values indicating better fit to the data i.e., sample size adjusted Bayesian Information Criterion (SSABIC) (Sclove, 1987), Akaike's Information Criteria (AIC) (Akaike, 1987), Bayesian Information Criterion (BIC) (Schwarz, 1978); ii) likelihood ratio statistical test methods comparing the model with K classes to a model with K-1 classes i.e., Lo-Mendell-Rubin likelihood ratio test (LRT) (Lo, Mendell, & Rubin, 2001), bootstrap likelihood ratio test (BLRT) (McCutcheon, 1987), and iii) entropy-based criterion goodness-of-fit indices based on the uncertainty of classification, ranging from 0 to 1 with a high score indicating good fit (entropy) (Jedidi, Ramaswamy, & Desarbo, 1993). We repeated the estimation procedure while varying the amount of missing data.

*Genetic data*

ALSPAC children were genotyped using the Illumina HumanHap550 quad chip genotyping platforms by 23andme subcontracting the Wellcome Trust Sanger Institute, Cambridge, UK and the Laboratory Corporation of America, Burlington, NC, US. The resulting raw genome-wide data were subjected to standard quality control methods. Individuals were excluded on the basis of gender mismatches; minimal or excessive heterozygosity; disproportionate levels of individual missingness (>3%) and insufficient sample replication (IBD < 0.8). Population stratification was assessed by multidimensional scaling analysis and compared with HapMap II (release 22) European descent (CEU), Han Chinese, Japanese and Yoruba reference populations; all individuals with non-European ancestry were removed. SNPs with a minor allele frequency of < 1%, a call rate of < 95% or evidence for violations of Hardy-Weinberg equilibrium ($p < 5 \times 10^{-7}$) were removed. Cryptic relatedness was measured as proportion of identity by descent (IBD > 0.1). Related subjects that passed all other quality control thresholds were retained during subsequent phasing and imputation. 9,115 subjects and 500,527 SNPs passed these quality control filters. ALSPAC mothers were also genotyped following a similar procedure, details of which are reported elsewhere (Taylor et al., 2018).

We combined 477,482 SNP genotypes in common between the sample of mothers and sample of children. We removed SNPs with genotype missingness above 1% due to poor quality (11,396 SNPs removed) and removed a further 321 subjects due to potential ID mismatches. This resulted in a dataset of 17,842 subjects containing 6,305 duos and 465,740 SNPs (112 were removed during liftover and 234 were out of HWE after combination). We estimated haplotypes using ShapeIT (v2.r644) which utilises relatedness during phasing. We obtained a phased version of the 1000 genomes reference panel (Phase 1, Version 3) from the Impute2 reference data repository (phased using ShapeIT v2.r644, haplotype release date Dec 2013). Imputation of the target data was performed using Impute V2.2.2 against the reference panel (all polymorphic SNPs excluding singletons), using all 2186 reference haplotypes (including non-Europeans). This resulted in 8,237 eligible ALSPAC children with available genotype data after exclusion of related subjects using cryptic relatedness measures described previously.

*Genetic Analyses*

Genome-wide association studies (GWAS) were conducted for each cognitive measure (working memory, emotion recognition and response inhibition) using all ALSPAC participants who completed the cognitive assessments and had available genetic data ($n$ = 2,471, $n$ = 2 ,560, and $n$ = 2,446, respectively). The same cognitive outcomes as in the observational analyses were used. Linear regression was conducted using SNPtest.2.5.0 to test associations between each single nucleotide polymorphism (SNP) and each cognitive phenotype under an additive model, controlling for age, sex, and the first 10 genetic principal components (to account for population stratification). Phenotypes were quantile normalized (using SNPtest) prior to analysis. Quality control checks were conducted on the summary data. SNPs were excluded if they deviated from Hardy-Weinberg equilibrium (at $p < 5x10^{-7}$), info of < 80%, and/or a minor allele frequency of < 1%. SNPs reaching $p < 5x10^{-8}$ were considered genome-wide significant. SNPs were then clumped to ensure independence at linkage disequilibrium (LD) $r^2 = 0.001$ and a distance of 10,000 kb, using the "clump_data" command in the TwoSampleMR R package (Hemani et al., 2018).

Genome-wide association analyses (GWAS) were conducted using ALSPAC participants who completed cognitive assessments ($n$~2,500). The same cognitive measures used in the observational analyses were used: *d'* as a measure of working memory, SSRT as a measure of response inhibition, and total number of recognised emotions as a measure of emotion recognition. Linear regression was conducted using SNPtest.2.5.0 to test associations between each single nucleotide polymorphism (SNP) and each cognitive phenotype under an additive model, controlling for age, sex, and the first 10 genetic principal components (to account for population stratification). SNPs reaching $p<5x10^{-8}$ were identified as genome-wide significant. Further details on the GWAS of cognitive functioning is provided by (Mahedy et al., 2019).

*Mendelian randomisation (MR)*

One-sample MR is the standard implementation of MR in a single data set with data on single nucleotide polymorphisms (SNPs), exposure, and outcome for all participants. Two-sample MR is an extension that allows for greater sample sizes resulting in greater statistical power. Contrary to a one-sample MR approach, in which the gene-exposure and gene-outcome relations are estimated in the same sample, two-sample approaches derive the estimates from separate samples (e.g., separate GWASs of exposure and outcome).

Two-sample MR was used to test the hypothesised causal effect of smoking initiation and lifetime cannabis use on cognitive functioning. The two-sample MR approach requires summary level data from GWAS, enabling SNP-outcome and SNP-exposure effects to be derived from different data sources. As the genetic instrument for smoking we used 378-independent genome-wide significant SNPs associated with smoking initiation identified by the GWAS & Sequencing Consortium of Alcohol and Nicotine use (GSCAN https://gscan.sph.umich.edu/) based on a sample of $N$~1,200,000. For cannabis use, we used 8-independent genome-wide significant SNPs associated with lifetime cannabis use based on the largest GWAS to date ($N$=184,765) (Pasman et al., 2018). As outcomes, we used GWAS conducted in ALSPAC ($n$~2,500) for each of our three primary outcome measures: i) working memory assessed using *d'*; ii) response inhibition assessed using SSRT; and iii) emotion recognition assessed using total number of correctly identified emotions (Mahedy et al., 2020). GWAS of working memory, response inhibition, and emotion recognition are available at the University of Bristol data repository, data.bris, (doi:10.5523).

Analyses were performed using the TwoSampleMR R package, part of MR-Base (Hemani et al., 2018). The inverse-variance weighted (IVW) approach was used as a primary analysis, with three complementary estimation methods as sensitivity analyses which each make different assumptions about the nature of horizontal pleiotropy (where the genetic variant associates with the outcome via an independent pathway to the exposure): MR Egger (Bowden, Davey Smith, & Burgess, 2015), weighted median (Bowden, Davey Smith, Haycock, & Burgess, 2016), and weighted mode (Hartwig, Davey Smith, & Bowden, 2017). A consistent effect across all of these methods would provide the most confidence that any observed effects are not due to pleiotropy.

The association between smoking initiation genetic score/ lifetime cannabis use and baseline confounders (gender, maternal smoking in pregnancy, housing tenure, maternal education, income, social position, head injury/unconsciousness before 11 years of age, working memory at 11 years of age, and alcohol use before age 14 years) were compared (Tables S9a and S9b). We found evidence on an association between our smoking initiation score and mothers who had >O level education, and alcohol use before 14 years of age. In terms of lifetime cannabis use, we found evidence of an association with head injury/ unconsciousness, working memory at age 11 years, and alcohol use before 14 years of age.

### RESULTS

#### *Two-sample MR*

Focusing on the IVW estimate as the primary measure, SNPs associated with smoking initiation were not associated with working memory (*b*=0.01 95%CI=-0.06 to 0.29; *p*=0.21); response inhibition (*b*=0.16 95%CI=-1.41 to 0.35; *p*=0.31); or emotion recognition (*b*=0.00 95%CI=-0.18 to 0.18; *p*=0.97). Other sensitivity estimates tended to be in the same direction and failed to demonstrate smoking initiation as a causal risk factor for deficits in cognitive functioning (Table S14a).

There was some evidence to suggest that SNPs associated with lifetime cannabis use was a causal risk factor for deficits in cognitive functioning (Table S14b). There was evidence of a possible causal association between lifetime cannabis use and poorer working memory using the IVW estimate (*b*=-0.37 95%CI=-0.72 to -0.02; *p*=0.04). Further sensitivity estimates (i.e., MR-Egger, weighted median, and weighted mode) were in the same direction providing some additional support for a possible causal association between SNPs associated with lifetime cannabis use and deficits in working memory. See Supplementary Material (Tables S14a and S14b). All other estimates failed to demonstrate cannabis use as a causal risk factor for deficits in cognitive functioning.

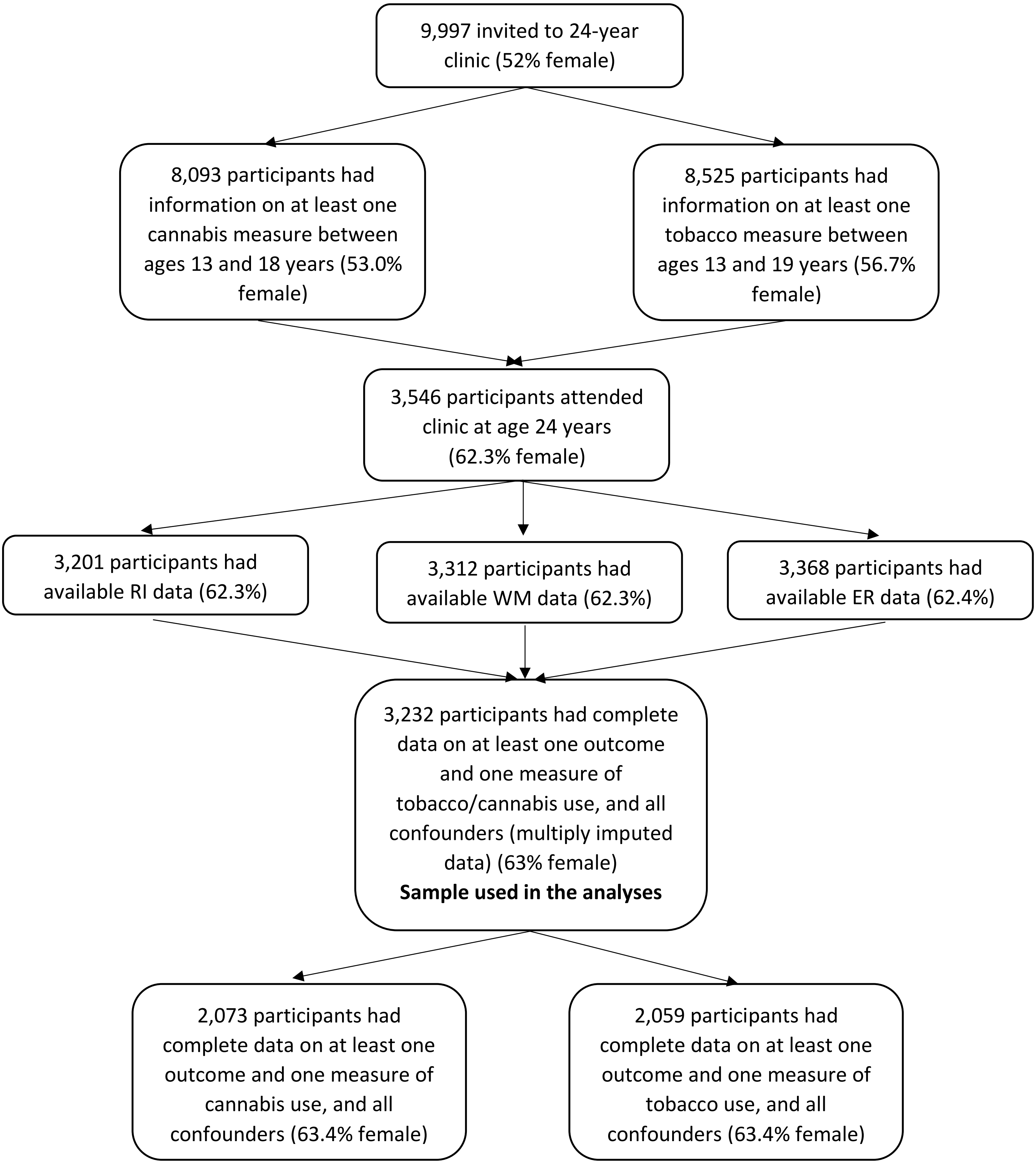
9,997 invited to 24-year clinic (52% female)
8,093 participants had information on at least one cannabis measure between ages 13 and 18 years (53.0% female)
8,525 participants had information on at least one tobacco measure between ages 13 and 19 years (56.7% female)
3,546 participants attended clinic at age 24 years (62.3% female)
3,201 participants had available RI data (62.3%)
3,312 participants had available WM data (62.3%)
3,368 participants had available ER data (62.4%)
3,232 participants had complete data on at least one outcome and one measure of tobacco/cannabis use, and all confounders (multiply imputed data) (63% female)
Sample used in the analyses
2,073 participants had complete data on at least one outcome and one measure of cannabis use, and all confounders (63.4% female)
2,059 participants had complete data on at least one outcome and one measure of tobacco use, and all confounders (63.4% female)

**Figure S1.** Sample attrition in ALSPAC.

Note: RI: response inhibition; WM: working memory; ER: emotion recognition

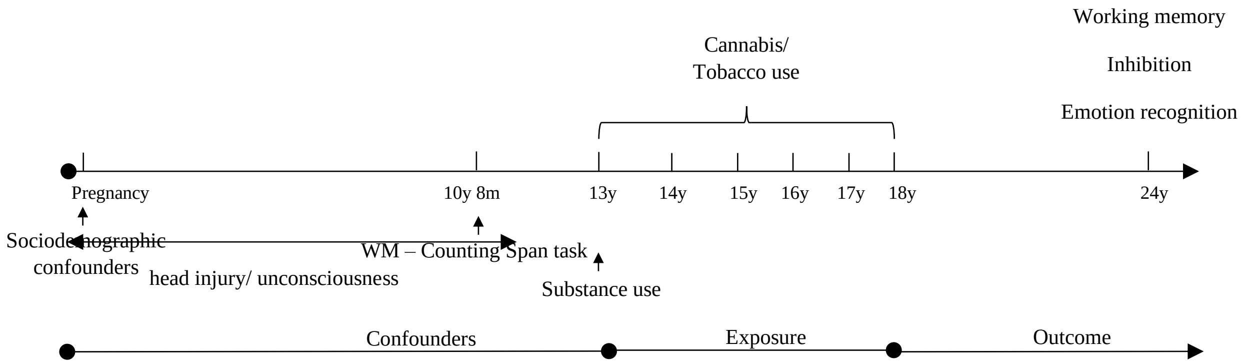


Figure S2. Timeline for data collection.

**Table S1a.** Comparison of model fit indices for tobacco use comparing 1 to 5 classes

| | # param | AIC | BIC | SSABIC | Entropy | Min class | LRT | BLRT |
|---|---|---|---|---|---|---|---|---|
| 1 class | 17 | 31904 | 32024 | 31970 | - | - | - | - |
| 2 class | 35 | 27447 | 27692 | 27581 | 0.76 | 22.5% | <0.0001 | <0.0001 |
| 3 class | 53 | 26668 | 27041 | 26873 | 0.69 | 11.9% | <0.0001 | <0.0001 |
| 4 class | 71 | 26485 | 26985 | 26760 | 0.72 | 3.4% | 0.22 | <0.0001 |
| 5 class | 89 | 26425 | 27052 | 26769 | 0.73 | 2.7% | 0.24 | <0.0001 |

Note: SSABIC: Bayesian Information Criterion; AIC: Akaike's Information Criteria; BIC: Bayesian Information Criterion; LRT: likelihood ratio test; BLRT: bootstrap likelihood ratio test

**Table S1b.** Comparison of model fit indices for cannabis use comparing 1 to 5 classes

| | # param | AIC | BIC | SSABIC | Entropy | Min class | LRT | BLRT |
|---|---|---|---|---|---|---|---|---|
| 1 class | | 18931 | 18931 | 18977 | - | - | - | - |
| 2 class | | 16551 | 16726 | 16646 | 0.77 | 14.4% | <0.0001 | <0.0001 |
| 3 class | | 16247 | 16513 | 16392 | 0.75 | 3.8% | <0.0001 | <0.0001 |
| 4 class | | 16119 | 16476 | 16476 | 0.77 | 2.9% | <0.0001 | <0.0001 |
| 5 class | | - | - | - | - | - | - | - |

Note: SSABIC: Bayesian Information Criterion; AIC: Akaike's Information Criteria; BIC: Bayesian Information Criterion; LRT: likelihood ratio test; BLRT: bootstrap likelihood ratio test; the 5-class model did not converge

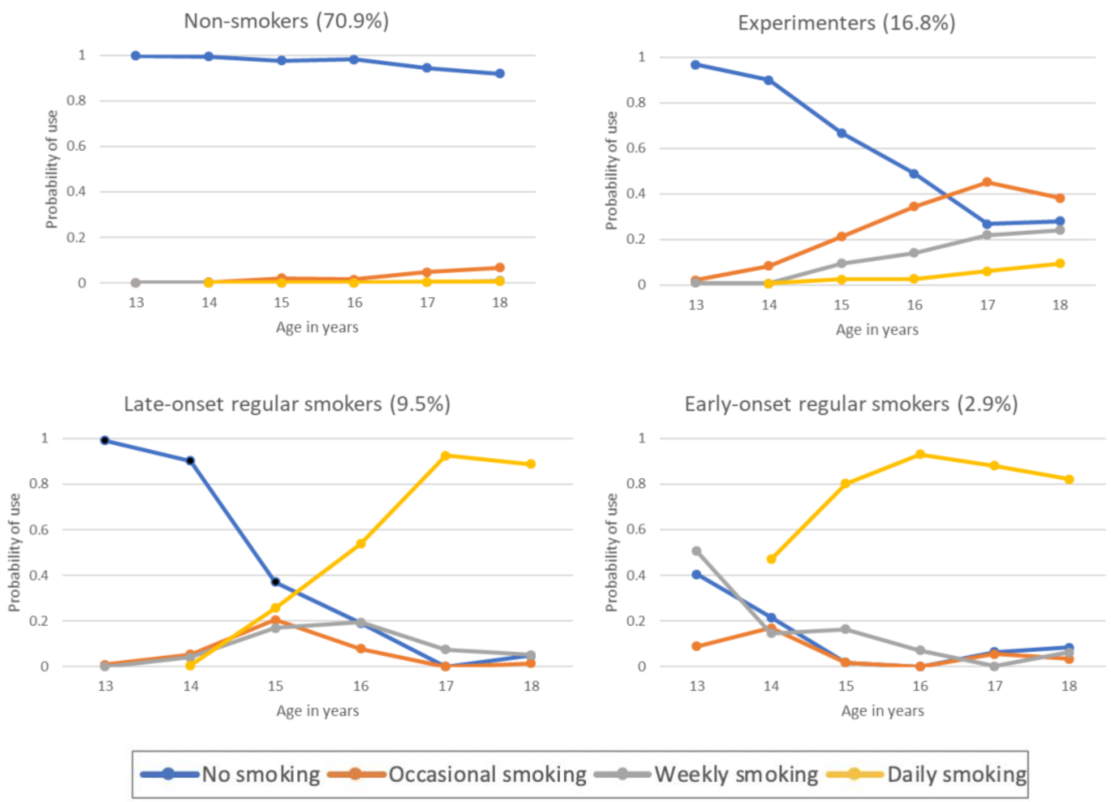


**Figure S3a.** Distribution of tobacco use across latent classes at each timepoint (*n*=8,525) [1]. Class proportions based on estimated posterior probability

[1] Tobacco use at age 13 was omitted from the figure as 'daily smoking' was not recorded at this age. Overall, the 'non-users' reported a low probability of tobacco use across all measurement occasions; 'experimenters' were mostly characterised by smoking on the later measurement occasions only; 'late-onset regular' smokers were mostly characterised by smoking on a daily basis in the later measurement occasions only; while 'early-onset regular' smokers were mostly characterised by smoking on a daily basis across timepoints from age 14 years onwards.

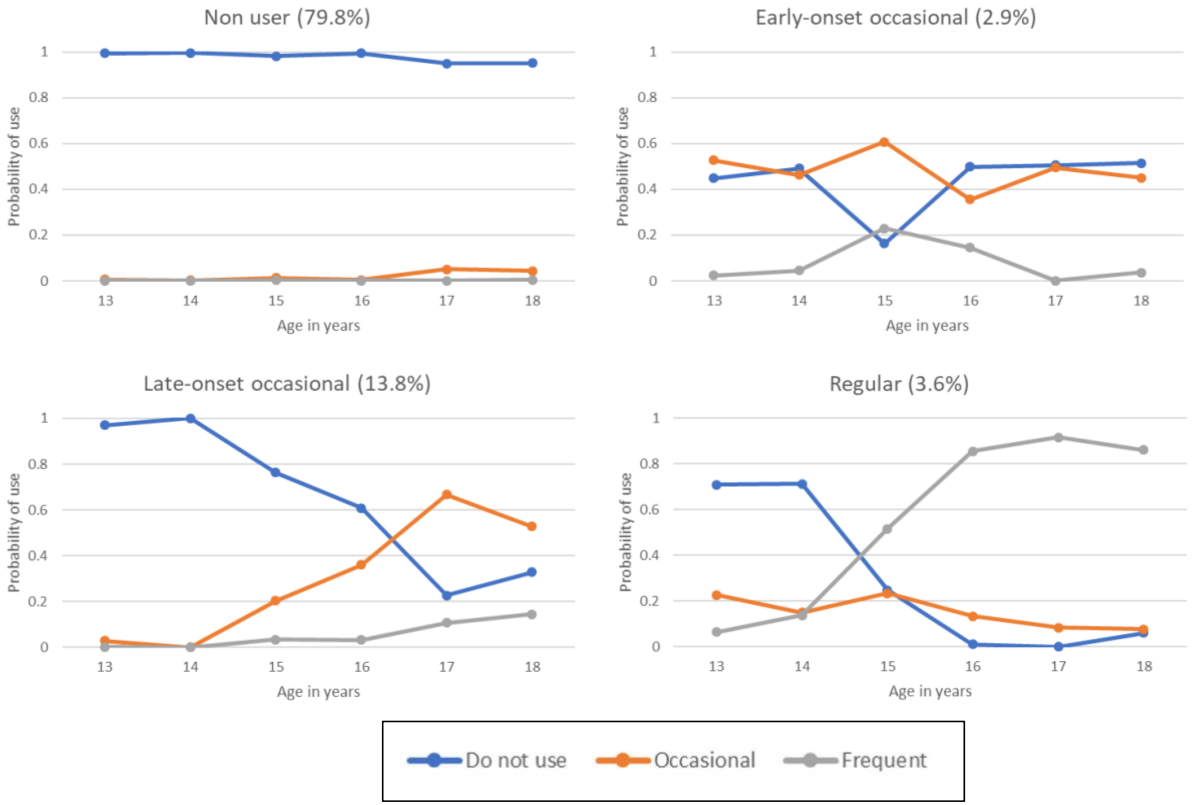


**Figure S3b.** Distribution of cannabis use across latent classes at each timepoint ($n$=8,093)[2]. Class proportions based on estimated posterior probability.

[2] The 'non users' reported a low probability of cannabis use across all measurement occasions; 'late-onset occasional' cannabis users were mostly characterised by cannabis use on the later measurement occasions only; 'early-onset occasional' cannabis users were mostly characterised by occasional cannabis use across all measurements; while 'regular cannabis users' were mostly characterised by cannabis use across timepoints with increasing involvement from age 15 years onwards.

**Table S2a.** Prevalence of tobacco use at each timepoint estimated using all available data

| | 13 | % | 14 | % | 15 | % | 16 | % | 17 | % | 18 | % |
|---|---|---|---|---|---|---|---|---|---|---|---|---|
| | 6,115 | | 5,926 | | 4,497 | | 5,066 | | 4,199 | | 3,334 | |
| Non-smoker | 5,964 | 97.5 | 5,579 | 94.1 | 3,558 | 79.1 | 4,070 | 80.3 | 3,019 | 71.9 | 2,438 | 73.1 |
| Occasional smoker | 47 | 0.8 | 154 | 2.6 | 381 | 8.5 | 369 | 7.3 | 461 | 11.0 | 357 | 10.7 |
| Weekly smoker | 104 | 1.7 | 75 | 1.3 | 202 | 4.5 | 215 | 4.2 | 195 | 4.6 | 156 | 4.7 |
| Daily smoker | - | - | 118 | 2.0 | 356 | 7.9 | 412 | 8.1 | 524 | 12.5 | 383 | 11.5 |

Note: Clinic assessments at ages 13, 15, and 17; questionnaires assessments at ages 14, 16, and 18

**Table S2b.** Prevalence of cannabis use at each timepoint estimated using all available data

| | 13 | % | 14 | % | 15 | % | 16 | % | 17 | % | 18 | % |
|---|---|---|---|---|---|---|---|---|---|---|---|---|
| | 5,786 | | 5,658 | | 5,060 | | 4,843 | | 3,915 | | 3,207 | |
| Non-user | 5,589 | 96.6 | 5,518 | 97.5 | 4,578 | 90.5 | 4,365 | 90.1 | 3,166 | 80.8 | 2,688 | 83.8 |
| Occasional use | 179 | 3.1 | 107 | 1.9 | 328 | 6.5 | 319 | 6.6 | 578 | 14.8 | 371 | 11.6 |
| Frequent use | 18 | 0.3 | 33 | 0.6 | 154 | 3.00 | 159 | 3.2 | 171 | 4.3 | 148 | 4.6 |

Note: Clinic assessments at ages 13, 15, and 17; questionnaires assessments at ages 14, 16, and 1

**Table S3.** Selective attrition for cognitive functioning assessed at the age 24 clinic

| | Available (*n*=3,201) *n* (%) | Not available (*n*=10,777) *n* (%) | OR (95% CI) |
|---|---|---|---|
| **Gender:** | | | |
| Males | 1,208 (37.7) | 6,009 (55.8) | 0.48 (0.44, 0.52) |
| **Income:** | | | |
| Low 20% | 359 (126) | 1,630 (23.1) | ref |
| 40% | 486 (17.0) | 1,480 (21.0) | 0.67 (0.58, 0.78) |
| 60% | 575 (20.1) | 1,401 (19.8) | 0.53 (0.46, 0.62) |
| 80% | 679 (23.7) | 1,306 (18.5) | 0.42 (0.37, 0.49) |
| Highest % | 760 (26.6) | 1,247 (17.7) | 0.36 (0.31, 0.42) |
| **Maternal education:** | | | |
| <O level | 1,559 (50.1) | 2,826 (30.4) | ref |
| O level | 1,040 (33.5) | 3,247 (35.0) | 1.72 (1.57, 1.89) |
| >O level | 509 (16.4) | 3,214 (34.6) | 3.48 (3.11, 3.90) |
| **Social**: | | | |
| i | 89 (3.0) | 593 (7.0) | ref |
| ii | 929 (30.1) | 3,547 (48.8) | 0.57 (0.45, 0.72) |
| iii | 1,379 (45.9) | 3,420 (40.3) | 0.37 (0.30, 0.47) |
| iv-v | 605 (20.2) | 919 (10.8) | 0.23 (0.18, 0.29) |
| **Tenure:** | | | |
| Mortgaged | 2,687 (86.4) | 6,853 (69.3) | ref |
| Private rent | 233 (7.5) | 1,152 (11.6) | 1.94 (1.67, 2.24) |
| Sub rent | 190 (6.1) | 1,890 (19.1) | 3.90 (3.34, 4.56) |
| **Maternal smoking:** | | | |
| Yes | 354 (12.0) | 2,172 (23.5) | 2.27 (2.00, 2.56) |
| **Head injury:** | | | |
| Yes | 106 (3.4) | 225 (3.4) | 0.98 (0.80, 1.21) |
| **Cigarette/cannabis:** | | | |
| None | 2,321 (87.0) | 2,212 (84.0) | ref |
| Smoking only | 178 (6.7) | 234 (8.9) | 1.38 (1.13, 1.69) |
| Smoking and cannabis | 168 (6.3) | 187 (7.1) | 1.17 (0.94, 1.45) |
| **WM at age 11:** | M (SD) | M (SD) | |
| Linear term | 3.51 (0.83) | 3.36 (0.86) | 0.82 (0.77, 0.86) |

Note: Maternal education: <O level indicating no qualification; O level: indicating completion of school examinations at age 16; and >O level: indicating completion of college or university education at or after age 18; SEP grouped into 4 categories: i—professional, ii—managerial, iii—skilled non-manual/skilled manual to iv-v—semi-skilled and unskilled occupations; lifetime cigarette smoking up to 16.5 years of age; lifetime cannabis use up to 16.5 years of age

**Table S4a**. Factors associated with tobacco use latent class membership

| | *N (%)* | Experimenters *b* (95% CI) | Late-onset regular *b* (95% CI) | Early-onset regular *b* (95% CI) | *Omnibus p* value |
|---|---|---|---|---|---|
| **Gender:** | | | | | |
| Males | 3,780 (46.9) | 0.78 (0.57, 0.98) | 0.35 (0.15, 0.55) | 0.86 (0.49, 1.24) | 0.09 |
| **Income:** | | | | | |
| Lowest 20% | 1,143 (16.3) | ref | ref | ref | <0.001 |
| 40% | 1,333 (19.0) | 0.38 (-0.09, 0.65) | -0.26 (-0.59, 0.06) | -0.35 (-0.83, 0.14) | |
| 60% | 1,428 (20.4) | -0.14 (-0.52, 0.25) | -0.43 (-0.75, -0.11) | -0.73 (-1.24, -0.21) | |
| 80% | 1,514 (21.6) | 0.35 (-0.00, 0.71) | -0.64 (-0.98, -0.30 | -1.01 (-1.58, -0.43) | |
| Highest | 1,589 (22.7) | 0.41 (0.06, 0.75) | -0.65 (-0.98, -0.32) | -2.56 (-3.88, -1.25) | |
| **Maternal education:** | | | | | |
| <O level | 3,247 (42.0) | ref | ref | ref | <0.001 |
| O level | 2,695 (34.9) | -0.26 (-0.47, -0.04) | 0.34 (0.10, 0.58) | 0.77 (0.25, 1.29) | |
| >O level | 1,776 (23.0) | -0.61 (-0.90, -0.32) | 0.48 (0.22, 0.74) | 1.42 (0.93, 1.92) | |
| **Social**: | | | | | |
| Semi/un-skilled | 322 (4.4) | ref | ref | ref | <0.001 |
| Skilled (non)manual | 2,576 (35.1) | -0.29 (-0.80, 0.23) | -0.22 (-0.74, 0.29) | -0.91 (-1.50, -0.31) | |
| Managerial | 3,269 (44.5) | -0.16 (-0.66, 0.35) | -0.30 (-0.81, 0.21) | -1.25 (-1.85, -0.65) | |
| Professional | 1,172 (16.0) | 0.07 (-0.45, 0.60) | -1.18 (-1.81, -0.54) | -2.48 (-3.60, -1.37) | |
| **Tenure:** | | | | | |
| Mortgaged | 6,369 (81.8) | ref | ref | ref | <0.001 |
| Sub rent | 651 (8.4) | 0.09 (-0.26, 0.44) | 0.55 (0.23, 0.86) | 0.63 (0.02, 1.24) | |
| Private rent | 762 (9.8) | -0.59 (-1.05, -0.12) | 0.58 (0.27, 0.89) | 1.67 (1.28, 2.05) | |
| **Maternal smoking:** | | | | | |
| Yes | 1,097 (14.9) | 0.03 (-0.29, 0.34) | 0.90 (0.65, 1.15) | 1.74 (1.37, 2.12) | <0.01 |
| **Head injury:** | | | | | |
| Yes | 269 (3.7) | 0.38 (-.10, .86) | 0.21 (-.35, .77) | 0.77 (0.06, 1.48) | 0.16 |
| **Ever alcohol use at age 13:** | | | | | |
| Yes | 1,848 (30.6) | 1.05 (0.82, 1.27) | 1.14 (0.90, 1.39) | 2.20 (1.74, 2.67) | 0.08 |

| | | | | | | |
|---|---|---|---|---|---|---|
| **WM at age 11:** | | M (SD) | | | | |
| | Linear term | 3.44 (0.8) | 1.05 (0.92, 1.19) | 0.82 (0.72, 0.94) | 0.73 (0.61, 0.87) | 0.11 |

**Table S4b**. Factors associated with cannabis use latent class membership

| | *N (%)* | Late-onset occasional<br>*b* (95% CI) | Early-onset occasional<br>*b* (95% CI) | Regular<br>*b* (95% CI) | *Omnibus*<br>*p* value |
|---|---|---|---|---|---|
| **Gender:** | | | | | |
| Males | 3,803 (47.0) | 0.09 (-0.13, 0.32) | -0.06 (-.49, .37) | -1.03 (-1.39, -0.66) | 0.03 |
| **Income:** | | | | | |
| Lowest 20% | 1,147 (16.3) | ref | ref | ref | <0.001 |
| 40% | 1,336 (19.0) | -0.13 (-0.55, 0.30) | 0.08 (-0.68, 0.85) | 0.07 (-0.45, 0.58) | |
| 60% | 1,431 (20.4) | 0.06 (-0.34, 0.45) | -0.31 (-1.16, 0.54) | -0.10 (-0.63, 0.42) | |
| 80% | 1,518 (21.6) | -0.09 (-0.50, 0.31) | 0.15 (-0.56, 0.87) | -0.92 (-1.62, -0.22) | |
| Highest | 1,592 (22.7) | 0.34 (-0.03, 0.72) | 0.11 (-0.64, 0.85) | -0.03 (-0.55, 0.49) | |
| **Maternal education:** | | | | | |
| <O level | 3,260 (42.1) | ref | ref | ref | <0.001 |
| O level | 2,702 (34.9) | -0.78 (-1.05, -0.51) | -0.48 (-0.99, 0.05) | -0.05 (-0.44, 0.35) | |
| >O level | 1,784 (23.0) | -0.98 (-1.34, -0.63) | -0.27 (-0.82, 0.28) | 0.23 (-0.17, 0.63) | |
| **Social**: | | | | | |
| Semi/un-skilled | 321 (4.4) | ref | ref | ref | <0.001 |
| Skilled (non)manual | 2,585 (35.1) | 0.28 (-0.59, 1.15) | -0.06 (-1.23, 1.13) | -0.63 (-1.23, -0.03) | |
| Managerial | 3,282 (44.5) | 0.86 (0.01, 1.71) | -0.08 (-1.27, 1.11) | -0.63 (-1.23, -0.03) | |
| Professional | 1,177 (16.0) | 1.02 (0.15, 1.89) | 0.37 (-0.85, 1.59) | -1.22 (-2.07, -0.37) | |
| **Tenure:** | | | | | |
| Mortgaged | 6,387 (81.8) | ref | ref | ref | <0.001 |
| Sub rent | 653 (8.4) | 0.12 (-0.27, 0.52) | 0.02 (-0.92, 0.95) | 0.85 (0.40, 1.30) | |
| Private rent | 764 (9.8) | -0.49 (-1.01, 0.04) | 0.80 (0.24, 1.35) | 0.85 (0.43, 1.27) | |
| **Maternal smoking:** | | | | | |
| Yes | 1,102 (14.9) | 0.15 (-0.18, 0.48) | 0.78 (0.26, 1.30) | 0.88 (0.52, 1.24) | 0.11 |
| **Head injury:** | | | | | |
| Yes | 273 (3.7) | 0.30 (-0.25, 0.85) | 0.30 (-0.82, 1.43) | 0.64 (-0.02, 1.30) | 0.27 |
| **Ever alcohol use at age 13:** | | | | | |
| Yes | 1,857 (30.7) | 0.63 (0.38, 0.88) | 1.29 (1.01, 1.56) | 1.06 (0.82, 1.29) | 0.41 |

| | | | | | | |
|---|---|---|---|---|---|---|
| **Tobacco at age 13** | | | | | | |
| | Yes | 737 (12.2) | 0.55 (0.12, 0.97) | 2.77 (2.28, 3.27) | 2.38 (1.99, 2.77) | <0.01 |
| **WM at age 11:** | | M (SD) | | | | |
| | Linear term | 3.44 (0.8) | 0.33 (0.17, 0.49) | -0.02 (-0.28, 0.24) | -0.32 (-0.57, -0.07) | 0.04 |

**Table S5** Sample characteristics who provided information on at least one of the cognitive measures at the 24 year clinic (3,380)

| | n (%) |
|---|---|
| **Gender** | |
| Female | 2,110 (62.4) |
| **Nicotine use** - Fagerstrom Test for Nicotine Dependence | |
| Very low dependence | 120 (3.6) |
| Low dependence | 249 (4.4) |
| Medium dependence | 45 (1.3) |
| High dependence | 56 (1.7) |
| Very high dependence | 16 (0.5) |
| **Cannabis use** – lifetime use | |
| >5 | 946 (6.0) |
| 5-20 | 543 (3.5) |
| 21-60 | 339 (2.2) |
| 61-100 | 154 (1.0) |
| 100+ | 432 (2.8) |
| **Alcohol -** AUDIT-C (cut-off ≥7) | |
| Yes | 988 (29.2) |
| **Depression** - Composite Interview Schedule – Revised (past year) | |
| Mild episode | 356 (10.5) |
| Moderate episode | 248 (7.3) |
| Severe episode | 39 (1.2) |
| **Anxiety** - Composite Interview Schedule – Revised (past year) | |
| Generalised Anxiety Disorder | 316 (9.4) |
| **Psychotic like symptoms -** symptom (ever) | |
| Yes | 104 (3.8) |
| **Medication** - Received for Mental Health problem (ever) | |
| Yes | 192 (1.2) |

**Table S6a.** Smoking patterns from 13 to 18 years and working memory at age 24 (lower *d′* reflect poorer performance)

| | No smoking | Experimenter | Late-onset regular | Early-onset regular | |
|---|---|---|---|---|---|
| *n*=3,232 for all models | Reference group | *b* (95% CI) | *b* (95% CI) | *b* (95% CI) | Wald (df) *p* value |
| **Unadjusted models** | | | | | |
| Working memory *d′* | - | 0.01 (-0.10, 0.10) | -0.32 (-0.48, -0.16) | -0.50 (-0.89, -0.10) | 28.92 (3) p<0.001 |
| **Adjusted for SES** | | | | | |
| Working memory *d′* | - | 0.00 (-0.11, 0.11) | -0.28 (-0.44, -0.12) | -0.39 (-0.78, 0.01) | 19.78 (3) p<0.001 |
| **Adjusted for SES/WM/HI** | | | | | |
| Working memory *d′* | - | 0.01 (-0.10, 0.11) | -0.27 (-0.43, -0.11) | -0.42 (-0.81, -0.02) | 20.25 (3) p<0.001 |
| **Fully adjusted models** | | | | | |
| Working memory *d′* | - | 0.01 (-0.12, 0.10) | -0.29 (-0.45, -0.13) | -0.45 (-0.84, -0.05) | 22.12 (3) p<0.001 |

Note. SES: socioeconomic status; WM: working memory at age ~11 years; HI: head injury/ unconsciousness up to age 11 years; Wald tests determine whether there were differences between patterns of tobacco use and subsequent cognitive functioning; negative estimates indicate poorer performance

**Table S6b.** Smoking patterns from 13 to 18 years and response inhibition at age 24 (longer reaction times reflect poorer performance)

| | No smoking | Experimenter | Late-onset regular | Early-onset regular | |
|---|---|---|---|---|---|
| *n*=3,232 for all models | Reference group | *b* (95% CI) | *b* (95% CI) | *b* (95% CI) | Wald (df) *p* value |
| **Unadjusted models** | | | | | |
| Stop signal reaction time | - | 0.02 (-0.05, 0.09) | 0.19 (-0.02, 0.40) | 0.21 (0.10, 0.31) | 22.73 (3) p<0.001 |
| **Adjusted for SES** | | | | | |
| Stop signal reaction time | - | 0.01 (-0.06, 0.09) | 0.08 (-0.13, 0.30) | 0.18 (0.07, 0.28) | 13.04 (3) p=0.005 |
| **Adjusted for SES/WM/HI** | | | | | |
| Stop signal reaction time | - | 0.01 (-0.06, 0.08) | 0.10 (-0.12, 0.32) | 0.17 (0.07, 0.27) | 13.05 (3) p=0.005 |
| **Fully adjusted model** | | | | | |
| Stop signal reaction time | - | 0.01 (-0.06, 0.09) | 0.10 (-0.12, 0.32) | 0.18 (0.07, 0.28) | 12.78 (3) p=0.005 |

Note. SES: socioeconomic status; WM: working memory at age ~11 years; HI: head injury/ unconsciousness up to age 11 years; Wald tests determine whether there were differences between patterns of tobacco use and subsequent cognitive functioning; positive estimates indicate poorer performance

**Table S6c.** Smoking patterns from 13 to 18 and emotion recognition at age 24 (lower scores reflect poorer performance)

| | No smoking | Experimenter | Late-onset regular | Early-onset regular | |
|---|---|---|---|---|---|
| *n*=3,232 for all models | Reference group | *b* (95% CI) | *b* (95% CI) | *b* (95% CI) | Wald (df) *p* value |
| **Unadjusted models** | | | | | |
| Total hits | - | 0.00 (-0.01, 0.01) | -0.02 (-0.04, -0.01) | -0.05 (-0.08, -0.01) | 20.32 (3) p<0.001 |
| **Adjusted for SES** | | | | | |
| Total hits | - | -0.00 (-0.01, 0.01) | -0.02 (-0.03, -0.00) | -0.04 (-0.07, -0.01) | 14.45 (3) p=0.002 |
| **Adjusted for SES/WM/HI** | | | | | |
| Total hits | - | -0.00 (-0.01, 0.01) | -0.02 (-0.03, -0.00) | -0.04 (-0.08, -0.01) | 14.65 (3) p=0.002 |
| **Fully adjusted model** | | | | | |
| Total hits | - | -0.00 (-0.01, 0.01) | -0.02 (-0.03, -0.00) | -0.04 (-0.08, -0.01) | 16.43 (3) p=0.001 |

Note. SES: socioeconomic status; WM: working memory at age ~11 years; HI: head injury/ unconsciousness up to age 11 years; Wald tests determine whether there were differences between patterns of tobacco use and subsequent cognitive functioning; negative estimates indicate poorer performance

**Table S7**. Tobacco use from 13 to 18 years and working memory indices at age 24 (lower scores reflect poorer performance)

| *n*=2,059 for all models | No smoking<br>Reference group | Experimenter<br>*b* (95% CI) | Late-onset regular<br>*b* (95% CI) | Early-onset regular<br>*b* (95% CI) | Wald (df) *p* value |
|---|---|---|---|---|---|
| **Unadjusted models** | | | | | |
| Number of hits | - | -0.93 (-9.56, 7.70) | -22.13 (-36.90, 7.63) | -53.47 (-80.54, -26.43) | 13.88 (3) p<0.01 |
| False alarms | | -0.00 (-0.02, 0.02) | 0.03 (-0.01, 0.08) | 0.06 (-0.04, 0.16) | 5.22 (3) p=0.15 |
| **Adjusted for SES** | | | | | |
| Number of hits | - | 0.01 (-0.01, 0.04) | -0.45 (-0.09, 0.00) | -0.13 (-0.26, 0.00) | 10.90 (3) p=0.01 |
| False alarms | | -0.00 (-0.02, 0.02) | 0.03 (-0.01, 0.07) | 0.05 (-0.05, 0.15) | 3.53 (3) p=0.31 |
| **Adjusted for SES/WM/HI** | | | | | |
| Number of hits | - | 0.01 (-0.01, 0.04) | -0.44 (-0.09, 0.00) | -0.13 (-0.27, -0.00) | 11.28 (3) p=0.01 |
| False alarms | | -0.00 (-0.02, 0.02) | 0.03 (-0.01, 0.07) | 0.06 (-0.05, 0.16) | 3.73 (3) p=0.29 |
| **Fully adjusted models** | | | | | |
| Number of hits | - | 0.01 (-0.01, 0.04) | -0.44 (-0.09, 0.00) | -0.13 (-0.27, -0.00) | 11.11 (3) p=0.01 |
| False alarms | | -0.02 (-0.02, 0.02) | 0.03 (-0.01, 0.07) | 0.06 (-0.05, 0.16) | 3.83 (3) p=0.28 |

**Table S8**. Tobacco use from 13 to 18 years and response inhibition indices at age 24 (longer reaction times reflect poorer performance)

| *n*=2,059 for all models | No smoking<br>Reference group | Experimenter<br>*b* (95% CI) | Late-onset regular<br>*b* (95% CI) | Early-onset regular<br>*b* (95% CI) | Wald (df) *p* value |
|---|---|---|---|---|---|
| **Unadjusted models** | | | | | |
| Go reaction time | - | 1.74 (-3.93, 7.42) | -1.34 (-10.90, 8.22) | 14.51 (-1.36, 30.37) | 3.45 (3) p=0.33 |
| Go accuracy | | -0.00 (0.01, 0.01) | -0.03 (-0.05, -0.01) | -0.06 (-0.11, 0.02) | 22.12 (3) p=0.0001 |
| Stop accuracy | | 0.00 (-0.02, 0.02) | -0.06 (-0.10, -0.02) | -0.13 (-0.22, 0.05) | 24.52 (3) p<0.0001 |
| **Adjusted for SES** | | | | | |
| Go reaction time | - | 1.16 (-4.55, 6.87) | -2.42 (-11.97, 7.14) | 11.00 (6.30, 28.29) | 1.73 (3) p=0.63 |
| Go accuracy | | -0.00 (-0.01, 0.01) | -0.03 (-0.04, -0.01) | -0.06 (-0.11, 0.01) | 18.88 (3) p<0.001 |
| Stop accuracy | | 0.00 (-0.02, 0.02) | -0.05 (-0.09, -0.02) | -0.12 (-0.21, 0.03) | 19.51 (3) p<0.001 |
| **Adjusted for SES/WM/HI** | | | | | |
| Go reaction time | - | 1.19 (-4.53, 6.92) | -2.53 (-12.08, 7.02) | 11.66 (-5.63, 28.96) | 1.96 (3) p=0.58 |
| Go accuracy | | -0.00 (-0.01, 0.01) | -0.03 (-0.04, -0.01) | -0.06 (-0.11, 0.02) | 19.26 (3) p<0.001 |
| Stop accuracy | | 0.00 (-0.02, 0.02) | -0.05 (-0.09, -0.02) | -0.12 (-0.21, 0.04) | 19.96 (3) p<0.001 |
| **Fully adjusted models** | | | | | |
| Go reaction time | - | 1.13 (-4.77, 7.03) | -2.63 (-12.35, 7.09) | 11.53 (-5.83, 28.90) | 1.89 (3) p=0.60 |
| Go accuracy | | -0.00 (-0.01, 0.01) | -0.03 (-0.04, -0.01) | -0.06 (-0.11, 0.01) | 17.30 (3) p<0.001 |
| Stop accuracy | | 0.00 (-0.02, 0.02) | -0.05 (-0.09, -0.02) | -0.13 (-0.21, 0.04) | 20.06 (3) p<0.001 |

**Table S9a.** Tobacco use from 13 to 18 years and specific emotion sensitivity at age 24 (lower scores reflect poorer performance)

| **Unadjusted models** *N*=2,059 | Low risk Reference group | Experimenters *b* (95% CI) | Late-onset regular *b* (95% CI) | Early-onset regular *b* (95% CI) | Wald (df) p value |
|---|---|---|---|---|---|
| Anger | - | 0.01 (-.02, .03) | -0.01 (-.06, .03) | -0.10 (-.20, .01) | 4.71 (3) p=0.19 |
| Disgust | - | 0.02 (-.01, .04) | -0.04 (-.08, .01) | -0.06 (-.17, .05) | 6.43 (3) p=0.09 |
| Fear | - | 0.00 (-.04, .03) | -0.08 (-.13, -.03) | -0.01 (-.15, .12) | 9.35 (3) p=0.02 |
| Happy | - | -0.01 (-.03, .01) | -0.03 (-.06, .00) | -0.02 (-.08, .04) | 5.00 (3) p=0.17 |
| Sad | - | -0.01 (-.03, .01) | -0.04 (-.07, -.01) | -0.06 (-.12, .00) | 11.50 (3) p=0.01 |
| Surprise | - | -0.01 (-.02, .01) | -0.05 (-.08, -.02) | 0.01 (-.05, .08) | 11.69 (3) p=0.01 |

**Table S9b.** Tobacco use from 13 to 18 years and specific emotion sensitivity at age 24 (lower scores reflect poorer performance)

| **Models adjusted for SES**<br>*N*=2,059 | Low risk<br>Reference group | Experimenters<br>*b* (95% CI) | Late-onset regular<br>*b* (95% CI) | Early-onset regular<br>*b* (95% CI) | Wald (df) p value |
|---|---|---|---|---|---|
| Anger | - | 0.01 (-.02, .03) | 0.00 (-.05, .04) | -0.09 (-.19, .02) | 3.00 (3) p=0.39 |
| Disgust | - | 0.01 (-.01, .04) | -0.03 (-.07, .01) | -0.06 (-.17, .06) | 4.60 (3) p=0.20 |
| Fear | - | 0.01 (-.04, .03) | -0.07 (-.12, -.01) | 0.00 (-.13, .13) | 6.47 (3) p=0.09 |
| Happy | - | -0.01 (-.03, .01) | -0.03 (-.06, .00) | -0.03 (-.09, .04) | 5.30 (3) p=0.15 |
| Sad | - | -0.01 (-.04, .01) | -0.03 (-.06, .00) | -0.06 (-.12, -.00) | 11.11 (3) p=0.01 |
| Surprise | - | -0.01 (-.03, .01) | -0.05 (-.07, -.02) | 0.02 (-.05, .08) | 10.03 (3) p=0.02 |

**Table S9c.** Tobacco use from 13 to 18 years and specific emotion sensitivity at age 24 (lower scores reflect poorer performance)

| **adjusted for SES/WM/HI**<br>*N*=2,059 | Low risk<br>Reference group | Experimenters<br>*b* (95% CI) | Late-onset regular<br>*b* (95% CI) | Early-onset regular<br>*b* (95% CI) | Wald (df) p value |
|---|---|---|---|---|---|
| Anger | - | 0.01 (-.02, .03) | 0.00 (-.05, .04) | -0.09 (-.20, .02) | 3.14 (3) p=0.37 |
| Disgust | - | 0.01 (-.01, .04) | -0.03 (-.07, .01) | -0.06 (-.17, .05) | 4.79 (3) p=0.19 |
| Fear | - | -0.01 (-.04, .03) | -0.06 (-.12, -.01) | -0.01 (-.14, .13) | 6.40 (3) p=0.09 |
| Happy | - | -0.01 (-.03, .01) | -0.03 (-.06, .00) | -0.03 (-.09, .04) | 5.38 (3) p=0.15 |
| Sad | - | -0.01 (-.04, .01) | -0.03 (-.06, .00) | -0.07 (-.13, -.00) | 11.26 (3) p=0.01 |
| Surprise | - | -0.01 (-.03, .01) | -0.04 (-.07, -.02) | 0.02 (-.05, .08) | 9.85 (3) p=0.02 |

**Table S9d.** Tobacco use from 13 to 18 years and specific emotion sensitivity at age 24 (lower scores reflect poorer performance)

| **Fully adjusted models**<br>*N*=2,059 | Low risk<br>Reference group | Experimenters<br>*b* (95% CI) | Late-onset regular<br>*b* (95% CI) | Early-onset regular<br>*b* (95% CI) | Wald (df) p value |
|---|---|---|---|---|---|
| Anger | - | 0.01 (-.02, .03) | 0.00 (-.05, .05) | -0.09 (-.20, .02) | 3.24 (3) p=0.36 |
| Disgust | - | 0.01 (-.02, .04) | -0.03 (-.08, .01) | -0.07 (-.19, .04) | 5.72 (3) p=0.12 |
| Fear | - | -0.01 (-.05, .03) | -0.07 (-.12, -.01) | -0.01 (-.14, .13) | 5.91 (3) p=0.12 |
| Happy | - | -0.01 (-.03, .01) | -0.03 (-.06, .00) | -0.03 (-.10, .03) | 5.72 (3) p=0.13 |
| Sad | - | -0.01 (-.04, .01) | -0.03 (-.06, .00) | -0.07 (-.13, -.00) | 16.09 (3) p=0.001 |
| Surprise | - | -0.01 (-.03, .01) | -0.04 (-.07, -.02) | 0.02 (-.05, .08) | 9.21 (3) p=0.03 |

**Table 10a.** Patterns of cannabis use from 13 to 18 years and working memory at age 24 (lower *d'* reflect poorer performance)

| | Non-user | Late-onset occasional | Early-onset occasional | Early-onset regular | |
|---|---|---|---|---|---|
| *n*=3,232 for all models | Reference group | *b* (95% CI) | *b* (95% CI) | *b* (95% CI) | Wald (df) *p* value |
| **Unadjusted models** | | | | | |
| Working memory *d'* | - | -0.01 (-0.13, 0.12) | 0.24 (-0.05, 0.52) | -0.58 (-0.90, -0.26) | 14.10 (3) p=0.003 |
| **Adjusted for SES** | | | | | |
| Working memory *d'* | - | -0.06 (-0.18, 0.07) | 0.17 (-0.11, 0.45) | -0.60 (-0.91, -0.30) | 16.26 (3) p<0.001 |
| **Adjusted for SES/WM/HI** | | | | | |
| Working memory *d'* | - | -0.09 (-0.21, 0.04) | 0.15 (-0.12, 0.43) | -0.60 (-0.91, -0.30) | 17.64 (3) p<0.001 |
| **Fully adjusted models** | | | | | |
| Working memory *d'* | - | -0.10 (-0.22, 0.03) | 0.12 (-0.17, 0.41) | -0.62 (-0.93, -0.31) | 18.56 (3) p<0.001 |

Note. SES: socioeconomic status; WM: working memory at age ~11 years; HI: head injury/ unconsciousness up to age 11 years; Wald tests determine whether there were differences between patterns of tobacco use and subsequent cognitive functioning; negative estimates indicate poorer performance

**Table 10b.** Patterns of cannabis use from 13 to 18 years and response inhibition at age 24 (longer reaction times reflect poorer performance)

| | Non-user | Late-onset occasional | Early-onset occasional | Early-onset regular | |
|---|---|---|---|---|---|
| *n*=3,232 for all models | Reference group | *b* (95% CI) | *b* (95% CI) | *b* (95% CI) | Wald (df) *p* value |
| **Unadjusted models** | | | | | |
| Stop signal reaction time | - | 0.00 (-0.07, 0.08) | 0.03 (-0.13, 0.18) | 0.29 (0.06, 0.52) | 7.02 (3) p=0.07 |
| **Adjusted for SES** | | | | | |
| Stop signal reaction time | - | 0.02 (-0.06, 0.10) | 0.04 (-0.13, 0.20) | 0.30 (0.07, 0.52) | 8.03 (3) p=0.04 |
| **Adjusted for SES/WM/HI** | | | | | |
| Stop signal reaction time | - | 0.04 (-0.04, 0.11) | 0.04 (-0.12, 0.21) | 0.30 (0.08, 0.52) | 9.25 (3) p=0.02 |
| **Fully adjusted model** | | | | | |
| Stop signal reaction time | - | 0.04 (-0.04, 0.11) | 0.05 (-0.11, 0.22) | 0.30 (0.08, 0.52) | 9.24 (3) p=0.02 |

Note. SES: socioeconomic status; WM: working memory at age ~11 years; HI: head injury/ unconsciousness up to age 11 years; Wald tests determine whether there were differences between patterns of tobacco use and subsequent cognitive functioning; negative estimates indicate poorer performance

**Table 10c.** Patterns of cannabis use from 13 to 18 and emotion recognition at age 24 (lower scores reflect poorer performance)

| | Non-user | Late-onset occasional | Early-onset occasional | Early-onset regular | |
|---|---|---|---|---|---|
| *n*=3,232 for all models | Reference group | *b* (95% CI) | *b* (95% CI) | *b* (95% CI) | Wald (df) *p* value |
| **Unadjusted models** | | | | | |
| Total hits | - | 0.00 (-0.01, 0.02) | 0.03 (0.00, 0.06) | -0.03 (-0.06, 0.00) | 7.71 (3) p=0.05 |
| **Adjusted for SES** | | | | | |
| Total hits | - | 0.00 (-0.01, 0.01) | 0.03 (0.00, 0.05) | -0.02 (-0.05, 0.01) | 4.68 (3) p=0.20 |
| **Adjusted for SES/WM/HI** | | | | | |
| Total hits | - | -0.00 (-0.01, 0.01) | 0.03 (-0.00, 0.05) | -0.02 (-0.05, 0.01) | 4.50 (3) p=0.21 |
| **Fully adjusted model** | | | | | |
| Total hits | - | -0.00 (-0.01, 0.01) | 0.02 (-0.00, 0.05) | -0.02 (-0.05, 0.01) | 4.23 (3) p=0.24 |

Note. SES: socioeconomic status; WM: working memory at age ~11 years; HI: head injury/ unconsciousness up to age 11 years; Wald tests determine whether there were differences between patterns of tobacco use and subsequent cognitive functioning; negative estimates indicate poorer performance

**Table S11**. Cannabis use from 13 to 18 years and working memory indices at age 24 (high scores reflect better performance)

| *n*=2,073 for all models | No smoking Reference group | Experimenter *b* (95% CI) | Late-onset regular *b* (95% CI) | Early-onset regular *b* (95% CI) | Wald (df) *p* value |
|---|---|---|---|---|---|
| **Unadjusted models** | | | | | |
| Number of hits | - | 0.00 (-0.03, 0.03) | 0.08 (0.01, 0.14) | -0.08 (-0.18, 0.01) | 7.45 (3) p=0.06 |
| False alarms | | 0.00 (-0.03, 0.02) | -0.03 (-0.06, -0.01) | 0.08 (-0.01, 0.18) | 9.22 (3) p=0.03 |
| **Adjusted for SES** | | | | | |
| Number of hits | - | 0.01 (-0.04, 0.02) | 0.07 (0.01, 0.13) | -0.09 (-0.18, 0.01) | 6.90 (3) p=0.07 |
| False alarms | | 0.00 (-0.02, 0.03) | -0.03 (-0.05, -0.00) | 0.08 (-0.02, 0.17) | 5.75 (3) p=0.13 |
| **Adjusted for SES/WM/HI** | | | | | |
| Number of hits | - | 0.02 (-0.05, 0.01) | 0.06 (-0.00, 0.13) | -0.09 (-0.18, 0.01) | 7.11 (3) p=0.07 |
| False alarms | | 0.00 (-0.02, 0.03) | -0.02 (-0.05, 0.00) | 0.08 (-0.02, 0.17) | 5.32 (3) p=0.15 |
| **Fully adjusted models** | | | | | |
| Number of hits | - | 0.02 (-0.05, 0.02) | 0.07 (-0.01, 0.14) | -0.09 (-0.18, 0.01) | 7.07 (3) p=0.07 |
| False alarms | | 0.01 (-0.02, 0.03) | -0.02 (-0.05, 0.00) | 0.08 (-0.02, 0.17) | 4.97 (3) p=0.17 |

**Table S12**. Cannabis use from 13 to 18 years and response inhibition indices at age 24 (longer reaction times reflect poorer performance)

| *n*=2,073 for all models | No smoking Reference group | Experimenter *b* (95% CI) | Late-onset regular *b* (95% CI) | Early-onset regular *b* (95% CI) | Wald (df) *p* value |
|---|---|---|---|---|---|
| **Unadjusted models** | | | | | |
| Go reaction time | - | -0.75 (-7.81, 6.30) | 1.26 (-14.91, 17.43) | -2.06 (-22.03, 17.92) | 0.10 (3) p=0.99 |
| Go accuracy | | -0.00 (-0.01, 0.01) | -0.02 (-0.02, 0.03) | -0.09 (-0.15, -0.03) | 8.73 (3) p=0.03 |
| Stop accuracy | | -0.02 (-0.04, 0.01) | -0.03 (-0.10, 0.04) | -0.14 (-0.23, -0.05) | 12.21 (3) p=0.003 |
| **Adjusted for SES** | | | | | |
| Go reaction time | - | 0.48 (-6.59, 7.55) | 1.11 (-15.40, 17.62) | 0.43 (-19.65, 20.50) | 0.05 (3) p=1.00 |
| Go accuracy | | -0.00 (-0.01, 0.01) | -0.01 (-0.02, 0.03) | -0.09 (-0.15, -0.03) | 8.80 (3) p=0.03 |
| Stop accuracy | | -0.02 (-0.04, 0.01) | -0.03 (-0.10, 0.04) | -0.14 (-0.23, -0.05) | 13.50 (3) p=0.004 |
| **Adjusted for SES/WM/HI** | | | | | |
| Go reaction time | - | 1.17 (-5.94, 8.29) | 1.53 (-15.05, 18.10) | 0.62 (-19.62, 20.85) | 0.16 (3) p=0.98 |
| Go accuracy | | -0.00 (-0.01, 0.01) | -0.01 (-0.02, 0.03) | -0.09 (-0.15, -0.03) | 9.82 (3) p=0.02 |
| Stop accuracy | | -0.02 (-0.04, 0.00) | -0.03 (-0.10, 0.04) | -0.14 (-0.22, -0.05) | 15.81 (3) p=0.001 |
| **Fully adjusted models** | | | | | |
| Go reaction time | - | 1.14 (-6.24, 8.52) | 1.46 (-15.57, 18.49) | 0.58 (-19.66, 20.81) | 0.13 (3) p=0.99 |
| Go accuracy | | -0.00 (-0.01, 0.01) | -0.01 (-0.02, 0.03) | -0.09 (-0.15, -0.03) | 9.17 (3) p=0.03 |
| Stop accuracy | | -0.02 (-0.05, 0.00) | -0.04 (-0.11, 0.04) | -0.14 (-0.23, -0.05) | 15.94 (3) p=0.001 |

**Table S13a.** Cannabis use from 13 to 18 years and specific emotion sensitivity at age 24 (lower scores reflect poorer performance)

| **Unadjusted models**<br>*N*=2,073 | Low risk<br>Reference group | Late-onset occasional<br>*b* (95% CI) | Early-onset occasional<br>*b* (95% CI) | Regular<br>*b* (95% CI) | Wald (df) p value |
|---|---|---|---|---|---|
| Anger | - | 0.03 (.00, .06) | 0.09 (.04, .16) | -0.09 (-.19, .01) | 14.66 (3) p<0.01 |
| Disgust | - | 0.03 (.00, .06) | 0.09 (.03, .16) | -0.08 (-.18, .02) | 12.12 (3) p<0.01 |
| Fear | - | 0.02 (-.02, .06) | 0.07 (-.04, .17) | -0.08 (-.17, .02) | 4.38 (3) p=0.22 |
| Happy | - | -0.03 (-.05, -.00) | -0.01 (-.07, .05) | -0.07 (-.14, .00) | 10.90 (3) p=0.01 |
| Sad | - | -0.00 (-.03, .02) | 0.01 (-.06, .09) | -0.06 (-.13, .01) | 3.10 (3) p=0.38 |
| Surprise | - | -0.00 (-.02, .02) | 0.02 (-.03, .07) | -0.08 (-.14, -.01) | 5.14 (3) p=0.16 |

**Table S13b.** Cannabis use from 13 to 18 years and specific emotion sensitivity at age 24 (lower scores reflect poorer performance)

| **Models adjusted for SES**<br>*N*=2,073 | Low risk<br>Reference group | Late-onset occasional<br>*b* (95% CI) | Early-onset occasional<br>*b* (95% CI) | Regular<br>*b* (95% CI) | Wald (df) p value |
|---|---|---|---|---|---|
| Anger | - | 0.03 (.00, .06) | 0.09 (.02, .15) | -0.07 (-.17, .02) | 12.97 (3) p<0.01 |
| Disgust | - | 0.03 (.00, .06) | 0.08 (.02, .15) | -0.06 (-.16, .04) | 9.66 (3) p=0.02 |
| Fear | - | 0.01 (-.03, .05) | 0.05 (-.05, .15) | -0.05 (-.14, .04) | 2.28 (3) p=0.52 |
| Happy | - | -0.03 (-.05, -.00) | -0.02 (-.07, .04) | -0.06 (-.12, .01) | 8.79 (3) p=0.03 |
| Sad | - | -0.00 (-.03, .02) | -0.00 (-.06, .07) | -0.04 (-.11, .02) | 1.85 (3) p=0.60 |
| Surprise | - | -0.00 (-.02, .02) | 0.02 (-.04, .07) | -0.06 (-.13, .00) | 3.62 (3) p=0.30 |

**Table S13c.** Cannabis use from 13 to 18 years and specific emotion sensitivity at age 24 (lower scores reflect poorer performance)

| **adjusted for SES/WM/HI**<br>*N*=2,073 | Low risk<br>Reference group | Late-onset occasional<br>*b* (95% CI) | Early-onset occasional<br>*b* (95% CI) | Regular<br>*b* (95% CI) | Wald (df) p value |
|---|---|---|---|---|---|
| Anger | - | 0.03 (.00, .06) | 0.08 (.02, .14) | -0.07 (-.17, .02) | 11.63 (3) p=0.01 |
| Disgust | - | 0.03 (.01, .06) | 0.08 (.01, .15) | -0.06 (-.16, .03) | 9.06 (3) p=0.03 |
| Fear | - | 0.01 (-.03, .05) | 0.05 (-.05, .15) | -0.05 (-.14, .04) | 1.93 (3) p=0.59 |
| Happy | - | -0.03 (-.05, -.00) | -0.02 (-.07, .04) | -0.06 (-.12, .01) | 9.36 (3) p=0.02 |
| Sad | - | -0.00 (-.03, .02) | -0.01 (-.07, .07) | -0.04 (-.11, .02) | 1.95 (3) p=0.58 |
| Surprise | - | -0.00 (-.03, .02) | 0.02 (-.04, .07) | -0.06 (-.12, .00) | 3.84 (3) p=0.28 |

**Table S13d.** Cannabis use from 13 to 18 years and specific emotion sensitivity at age 24 (lower scores reflect poorer performance)

| **Fully adjusted models**<br>*N*=2,073 | Low risk<br>Reference group | Late-onset occasional<br>*b* (95% CI) | Early-onset occasional<br>*b* (95% CI) | Regular<br>*b* (95% CI) | Wald (df) p value |
|---|---|---|---|---|---|
| Anger | - | 0.04 (.00, .07) | 0.09 (.03, .16) | -0.07 (-.16, .03) | 12.83 (3) p=0.01 |
| Disgust | - | 0.03 (.01, .06) | 0.08 (.01, .15) | -0.06 (-.16, .04) | 8.56 (3) p=0.04 |
| Fear | - | 0.01 (-.03, .05) | 0.05 (-.05, .16) | -0.05 (-.14, .04) | 2.02 (3) p=0.57 |
| Happy | - | -0.03 (-.05, -.00) | -0.02 (-.08, .04) | -0.06 (-.13, .01) | 9.13 (3) p=0.03 |
| Sad | - | -0.01 (-.03, .02) | -0.01 (-.08, .06) | -0.05 (-.11, .02) | 2.71 (3) p=0.44 |
| Surprise | - | -0.00 (-.02, .02) | 0.02 (-.04, .07) | -0.06 (-.12, .00) | 3.71 (3) p=0.29 |

**Table S14a.** Associations between genetic instrument for smoking initiation and confounders

| | | Smoking PRS | | Smoking observed | |
|---|---|---|---|---|---|
| | N | Estimate (95% CI) | *p* | Estimate (95% CI) | *p* |
| Gender | 2,444 | | | | |
| Female | 53.1% | ref | | | |
| Male | 46.9% | -108.8 (-344.3, 126.8) | 0.37 | - | |
| Mat smoking in preg | 2,284 | | | | |
| No | 85.1% | ref | | | |
| Yes | 14.9% | 348.9 (-26.1, 723.9) | 0.70 | - | |
| Tenure | 2,407 | | | | |
| Mortgaged | 81.8% | ref | | | |
| Subs | 8.4% | 119.2 (-327.5, 566.0) | 0.60 | - | |
| Priv | 9.8% | 145.7 (-385.2, 676.7) | 0.59 | - | |
| Mat educ | 2,401 | | | | |
| <O level | 50.1% | ref | | | |
| O level | 16.4% | 233.1 (-24.54, 490.8) | 0.08 | - | |
| >O level | 33.5% | 404.5 (60.8, 748.2) | 0.02 | 0.40 (0.10, 0.68) | 0.007 |
| Income | 2,219 | | | | |
| Lowest 20% | 16.3% | ref | | | |
| 21-40% | 19.0% | 44.3 (-420.1, 508.8) | 0.85 | - | |
| 41-60% | 20.4% | -197.2 (-644.7, 250.3) | 0.39 | - | |
| 61-80% | 21.6% | -93.9 (-524.6, 336.9) | 0.67 | - | |
| Highest 20% | 22.7% | -115.9 (-538.7, 306.9) | 0.59 | - | |
| Social | 2,322 | | | | |
| Semi/un-skilled | 4.4% | ref | | | |
| Skilled (non)manual | 35.1% | -54.40 (-815.8, 707.0) | 0.89 | - | |
| Managerial | 44.5% | 417.7 (77.7, 757.6) | 0.02 | 0.51 (0.19, 0.83) | 0.002 |
| Professional | 16.0% | 259.4 (-49.9, 568.8) | 0.10 | - | |
| Head injury | 2,386 | | | | |
| No | 96.6% | ref | | | |
| Yes | 3.4% | -288.0 (-906.7, 330.7) | 0.36 | - | |
| Working memory | 2,069 | -33.84 (-139.9, 72.3) | 0.53 | - | |
| Alcohol use < age 14 | 2,099 | | | | |
| No | 69.3% | ref | | | |
| Yes | 30.7% | 473.4 (219.6, 727.2) | <0.001 | 2.01 (1.74, 2.29) | <0.001 |

Note: observed observations are reported where there as evidence of an association between genetic score and confounders

**Table S14b.** Associations between genetic instrument for lifetime cannabis use and confounders

| | | Cannabis PRS | | Cannabis observed | |
|---|---|---|---|---|---|
| | N | Estimate (95% CI) | *p* | Estimate (95% CI) | *p* |
| Gender | 2,444 | | | | |
| Female | 53.1% | ref | | | |
| Male | 46.9% | -2.70 (-12.16, 6.75) | 0.58 | - | |
| Mat smoking in preg | 2,284 | | | | |
| No | 85.1% | ref | | | |
| Yes | 14.9% | -0.46 (-15.42, 14.50) | 0.95 | - | |
| Tenure | 2,407 | | | | |
| Mortgaged | 81.8% | ref | | | |
| Subs | 8.4% | -6.03 (-24.06, 11.99) | 0.51 | - | |
| Priv | 9.8% | 18.43 (-2.28, 39.15) | 0.08 | - | |
| Mat educ | 2,401 | | | | |
| <O level | 50.1% | ref | | | |
| O level | 16.4% | -3.13 (-13.42, 7.16) | 0.55 | - | |
| >O level | 33.5% | -0.21 (-13.89, 13.46) | 0.98 | - | |
| Income | 2,219 | | | | |
| Lowest 20% | 16.3% | ref | | | |
| 21-40% | 19.0% | 18.94 (0.10, 37.79) | 0.50 | - | |
| 41-60% | 20.4% | 13.18 (-5.04, 31.40) | 0.16 | - | |
| 61-80% | 21.6% | 15.03 (-2.52, 32.57) | 0.09 | - | |
| Highest 20% | 22.7% | 10.80 (-6.45, 28.06) | 0.22 | - | |
| Social | 2,322 | | | | |
| Semi/un-skilled | 4.4% | ref | | | |
| Skilled (non)manual | 35.1% | 15.88 (-13.80, 45.58) | 0.29 | - | |
| Managerial | 44.5% | -0.40 (-13.87, 13.08) | 0.95 | - | |
| Professional | 16.0% | -1.17 (-13.46, 11.12) | 0.85 | - | |
| Head injury | 2,386 | | | | |
| No | 96.6% | ref | | | |
| Yes | 3.4% | -25.94 (-51.83, -0.04) | 0.05 | 0.70 (-0.09, 1.50) | 0.08 |
| Working memory | 2,069 | -4.63 (-8.85, -0.40) | 0.03 | 0.11 (-0.07, 0.29) | 0.23 |
| Alcohol use < age 14 | 2,099 | | | | |
| No | 69.3% | ref | | | |
| Yes | 30.7% | 12.06 (2.06, 22.06) | 0.02 | 3.27 (2.27, 4.28) | <0.001 |

**Table S15a.** Two-sample MR analyses of the effects of smoking initiation on cognitive functioning (unstandardized coefficients)

| Exposure | Outcome | Method | N SNPs | Beta (95% CI) | P-value |
|---|---|---|---|---|---|
| **Smoking initiation** | **Working memory** | Inverse-Variance Weighted | 341 | 0.01 (-0.06, 0.29) | 0.21 |
| | | MR Egger (SIMEX) | 341 | -0.68 (-1.41, 0.06) | 0.07 |
| | | Weighted Median | 341 | 0.06 (-0.21, 0.32) | 0.65 |
| | | Weighted Mode | 341 | -0.32 (-1.11, 0.48) | 0.43 |
| **Smoking initiation** | **Response inhibition** | Inverse-Variance Weighted | 341 | 0.16 (-0.03, 0.35) | 0.10 |
| | | MR Egger (SIMEX) | 341 | 1.30 (-2.22, 0.03) | 0.02 |
| | | Weighted Median | 341 | 0.21 (-0.07, 0.49) | 0.14 |
| | | Weighted Mode | 341 | 0.30 (-0.55, 1.16) | 0.49 |
| **Smoking initiation** | **Emotion recognition** | Inverse-Variance Weighted | 341 | 0.00 (-0.18, 0.18) | 0.97 |
| | | MR Egger (SIMEX) | 341 | 0.04 (-0.71, 0.79) | 0.91 |
| | | Weighted Median | 341 | -0.05 (-0.31, 0.21) | 0.68 |
| | | Weighted Mode | 341 | -0.19 (-1.04, 0.67) | 0.67 |

**Table S15b.** Two-sample MR analyses of the effects of lifetime cannabis use on cognitive functioning (unstandardized coefficients)

| Exposure | Outcome | Method | N SNPs | Beta (95% CI) | P-value |
|---|---|---|---|---|---|
| **Lifetime cannabis use** | **Working memory** | Inverse-Variance Weighted | 8 | -0.37 (-0.72, -0.02) | 0.04 |
| | | MR Egger (SIMEX) | 8 | -0.10 (-0.21, 0.03) | 0.17 |
| | | Weighted Median | 8 | -0.38 (-0.81, 0.06) | 0.09 |
| | | Weighted Mode | 8 | -0.21 (-0.85, 0.43) | 0.55 |
| **Lifetime cannabis use** | **Response inhibition** | Inverse-Variance Weighted | 8 | 0.04 (-0.32, 0.39) | 0.85 |
| | | MR Egger (SIMEX) | 8 | 0.50 (-0.22, 0.02) | 0.32 |
| | | Weighted Median | 8 | 0.22 (-0.24, 0.68) | 0.35 |
| | | Weighted Mode | 8 | 0.26 (-0.35, 0.87) | 0.43 |
| **Lifetime cannabis use** | **Emotion recognition** | Inverse-Variance Weighted | 8 | -0.08 (-0.52, 0.36) | 0.71 |
| | | MR Egger (SIMEX) | 8 | 0.06 (-0.22, 0.03) | 0.93 |
| | | Weighted Median | 8 | 0.02 (-0.46, 0.48) | 0.95 |
| | | Weighted Mode | 8 | 0.01 (-0.67, 0.69) | 0.98 |

**Table S16.** Tests of the unweighted and weighted regression dilution $I^2_{GX}$

| | $I^2_{GX}$ **Unweighted** | $I^2_{GX}$ **Weighted** | **mF** |
|---|---|---|---|
| **Smoking** | | | |
| Smoking initiation > working memory | 0.608 | 0.390 | 44.88 |
| Smoking initiation > response inhibition | 0.608 | 0.390 | 44.88 |
| Smoking initiation > emotion recognition | 0.608 | 0.390 | 44.88 |
| **Cannabis** | | | |
| Cannabis lifetime > working memory | 0.704 | 0.288 | 38.65 |
| Cannabis lifetime > response inhibition | 0.704 | 0.286 | 38.65 |
| Cannabis lifetime > emotion recognition | 0.704 | 0.285 | 38.65 |

Note. Unweighted estimates only take into account dilution in the SNP-exposure effects, whereas weighted estimates account for the SE of the SNP-outcome effects (Bowden, Del Greco M, et al., 2016). mF is the mean F-statistic.

**Table S17.** Tests of heterogeneity in the SNP-exposure association using the IVW method

| | Cochran's Q | df | p-value |
|---|---|---|---|
| **Smoking** | | | |
| Smoking initiation > working memory | 346.42 | 340 | 0.39 |
| Smoking initiation > response inhibition | 400.56 | 340 | 0.01 |
| Smoking initiation > emotion recognition | 368.64 | 340 | 0.14 |
| **Cannabis** | | | |
| Cannabis lifetime > working memory | 2.49 | 6 | 0.87 |
| Cannabis lifetime > response inhibition | 5.11 | 6 | 0.53 |
| Cannabis lifetime > emotion recognition | 9.63 | 6 | 0.14 |
